# An Empirical Assessment of Inferential Reproducibility of Linear Regression in Health and Biomedical Research Papers

**DOI:** 10.64898/2026.04.07.26350296

**Authors:** Lee Jones, Adrian Barnett, Gunter Hartel, Dimitrios Vagenas

## Abstract

**Background:** In health research, variability in modelling decisions can lead to different conclusions even when the same data are analysed, a challenge known as inferential reproducibility. In linear regression analyses, incorrect handling of key assumptions, such as normality of the residuals and linearity, can undermine reproducibility. This study examines how violations of these assumptions influence inferential conclusions when the same data are reanalysed.

**Methods:** We randomly sampled 95 health-related *PLOS ONE* papers from 2019 that reported linear regression in their methods. Data were available for 43 papers, and 20 were assessed for computational reproducibility, with three models per paper evaluated. The 14 papers that included a model at least partially computationally reproduced were then examined for inferential reproducibility. To assess the impact of assumption violations, differences in coefficients, 95% confidence intervals, and model fit were compared.

**Results:** Of the fourteen papers assessed, only three were inferentially reproducible. The most frequently violated assumptions were normality and independence, each occurring in eight papers. Violations of independence were particularly consequential and were commonly associated with inferential failure. Although reproduced analyses often retained the same binary statistical significance classification as the original studies, confidence intervals were frequently wider, indicating greater uncertainty and reduced precision. Such uncertainty may affect the interpretation of results and, in turn, influence treatment decisions and clinical practice.

**Conclusion:** Our findings demonstrate that substantial violations of key modelling assumptions often went undetected by authors and peer reviewers and, in many cases, were associated with inferential reproducibility failure. This highlights the need for stronger statistical education and greater transparency in modelling decisions. Rather than applying rigid or misinformed rules, such as incorrectly testing the normality of the outcome variable, researchers should adopt modelling frameworks guided by the research question and the study design. When assumptions are violated, appropriate alternatives, such as robust methods, bootstrapping, generalised linear models, or mixed-effects models, should be considered. Given that assumption violations were common even in relatively simple regression models, early and sustained collaboration with statisticians is critical for supporting credible, defensible, and clinically meaningful conclusions.

## Introduction

The reproducibility of health research is essential for assessing the reliability of findings and supporting informed clinical decision-making. When research is reproducible, healthcare professionals can be more confident that their decisions are based on trustworthy evidence, thereby improving outcomes and reducing the risk of harm from ineffective or inappropriate interventions [1]. However, many published health and biomedical studies cannot be reliably reproduced, contributing to what is now widely recognised as a reproducibility crisis [2, 3].

Reproducibility concerns highlight the need to strengthen research standards through rigorous study design, appropriate statistical methods, and transparent reporting. Empirical assessments by Hardwicke et al. [4] and others have shown that essential materials, including data, code, and analysis plans, are frequently unavailable or insufficiently documented, limiting the ability to reproduce published findings [5]. These challenges raise concerns about the reliability of scientific evidence and emphasise the importance of reproducibility in supporting sound healthcare decisions and safe, effective patient care, enabling a more efficient use of limited resources on interventions more likely to benefit public health [6].

In this paper, we focus on inferential reproducibility, defined as the consistency of conclusions drawn from the same dataset when alternative, reasonable analytical choices are applied [3]. Although methodological and computational reproducibility have received increasing attention, inferential reproducibility remains comparatively underexplored [3]. Differences in analytical decisions, such as the choice of statistical test, covariate adjustment, model specification, or outcome prioritisation, may be legitimate in principle. However, when these decisions lack clear justification or transparency, they can lead to divergent interpretations of the same data [7, 8], undermining confidence in research findings and complicating the identification of effective treatments.

The reproducibility crisis is further exacerbated by limited transparency in how statistical conclusions are derived. Researchers may fail to document their analytical rationale or consider alternative approaches when assumptions are violated, and formal assessment of assumptions is often absent from publications [9]. Inconsistent statistical practices, gaps in training, and limited access to methodological expertise further contribute to variability in statistical inferences and conclusions. Publication incentives that prioritise novelty and statistical significance may also encourage selective reporting and over-interpretation [10].

These challenges are particularly relevant for linear regression, one of the most widely used statistical methods in health research [11]. The validity of linear regression relies on assumptions of linearity, independence, homoscedasticity, and normality of residuals. Violations of these assumptions can meaningfully alter effect estimates, standard errors, and statistical inference [12, 13]. This study examines how such violations contribute to variability in inferential reproducibility, in which analytical decisions can lead to differing conclusions despite using identical data. It provides a practical guide for assessing statistical assumptions and offers recommendations to improve transparency, promote rigour, and support more reliable evidenced based decision making.

## Materials and Methods

This study is the second part of a project examining the statistical quality of published research. In Stage 1, experienced statisticians reviewed selected papers for linear regression reporting practices, examining whether authors reported checking statistical assumptions and how those assumptions were assessed [14, 15]. Stage 2 examines reproducibility. The first component of Stage 2 assessed computational reproducibility, examining whether data were available and whether published results could be reproduced from the available materials [16]. This paper presents the second component of Stage 2, assessing inferential reproducibility, specifically, whether the reported conclusions are justified based on the statistical assumptions.

### Sample Size Determination

A sample of 100 papers was selected to evaluate the prevalence of reported linear regression assumptions. The sample size was calculated to estimate a proportion of 0.05 (5%) with a two-sided 95% confidence interval and a margin of error of 5% [14]. It was anticipated that approximately 50% of papers would contain accessible data, enabling reproducibility assessment. This study was designed as a pilot, given the current lack of empirical evidence on the proportion of published studies that are reproducible.

### Selection of Papers

The initial sample of 100 research articles, excluding editorials and other non-research content, was randomly selected from *PLOS ONE* publications in 2019 to provide a relevant snapshot of reporting practices. This time frame also offered an unintended advantage: it was the last full year prior to the COVID-19 pandemic, thereby avoiding potential disruptions to research [17]. *PLOS ONE* was selected because its long-established data-sharing policy required authors to make the data underlying their findings publicly available [18].

The selection used the ‘searchplos’ function from the ‘rplos’ R package [19] to identify articles that met two criteria: (1) the subject area included “health,” and (2) the Materials and Methods section reported the use of “linear regression.” Eligible papers were randomly ordered, and the first 100 meeting the inclusion criteria were selected. To focus on standard applications of linear regression in health research, articles were excluded if they used clustered or random effects, alternative modelling approaches (e.g., Bayesian or non-parametric methods), or applied linear regression only for secondary purposes (e.g., calibration or pre-processing). Upon full-text review, five papers were excluded as they did not report any linear regression results, leaving 95 papers for the assessment of reporting practices.

### Research Questions

- What proportion of analyses and papers violated statistical assumptions (independence, linearity, homoscedasticity, and normality of residuals), including issues with outliers or multicollinearity?
- How many papers reported that the authors had checked statistical assumptions, outliers, and multicollinearity?
- What proportion of analyses remained inferentially reproducible after accounting for unmet assumptions, outliers, and multicollinearity?

### Evaluating Model Assumptions and Reproducibility

Reproducibility analyses were conducted by the primary author, an accredited senior biostatistician with over 20 years of experience in statistical modelling and analysis of healthcare data. To ensure the reliability of the findings, selected results were independently reviewed by three senior biostatisticians, two of whom are accredited (AStat). Agreement on the reproduced results and interpretation was reached through discussion and consensus. The analyses followed a consistent, transparent process, documenting all steps and decisions. We refer to each paper by a unique number, the full list of identifiers and corresponding DOIs are available in the GitHub data [20].

A maximum of three linear regression analyses per paper was selected to ensure the workload remained feasible. Because models within a paper typically share the same data and analytical framework, their reproducibility was expected to be highly correlated, limiting the additional insight gained from reproducing all models. For papers with more than three models, the final or primary models of interest were included first, with any remaining models selected at random. If no primary model was identified, all three were selected at random. Models that were partially reproducible were further assessed for inferential reproducibility, using the reproduced results and available data to check model assumptions and conduct sensitivity analyses.

The assumptions of linear regression were evaluated using descriptive statistics, diagnostic plots, and formal tests, with additional checks for multicollinearity, outliers, and influential observations. A simulated example illustrating the relationship between blood pressure and age is shown in Fig 1 to demonstrate how these diagnostic procedures were applied in practice. While full implementation details are provided below and on GitHub [20], this example highlights key components of assumption checking, including inspection of scatter plots to assess linearity and potential non-linear (e.g., quadratic) relationships, examination of residual plots for evidence of non-linearity or heteroscedasticity, identification of influential observations, and assessment of whether residuals are approximately normally distributed.

**Figure 1.**
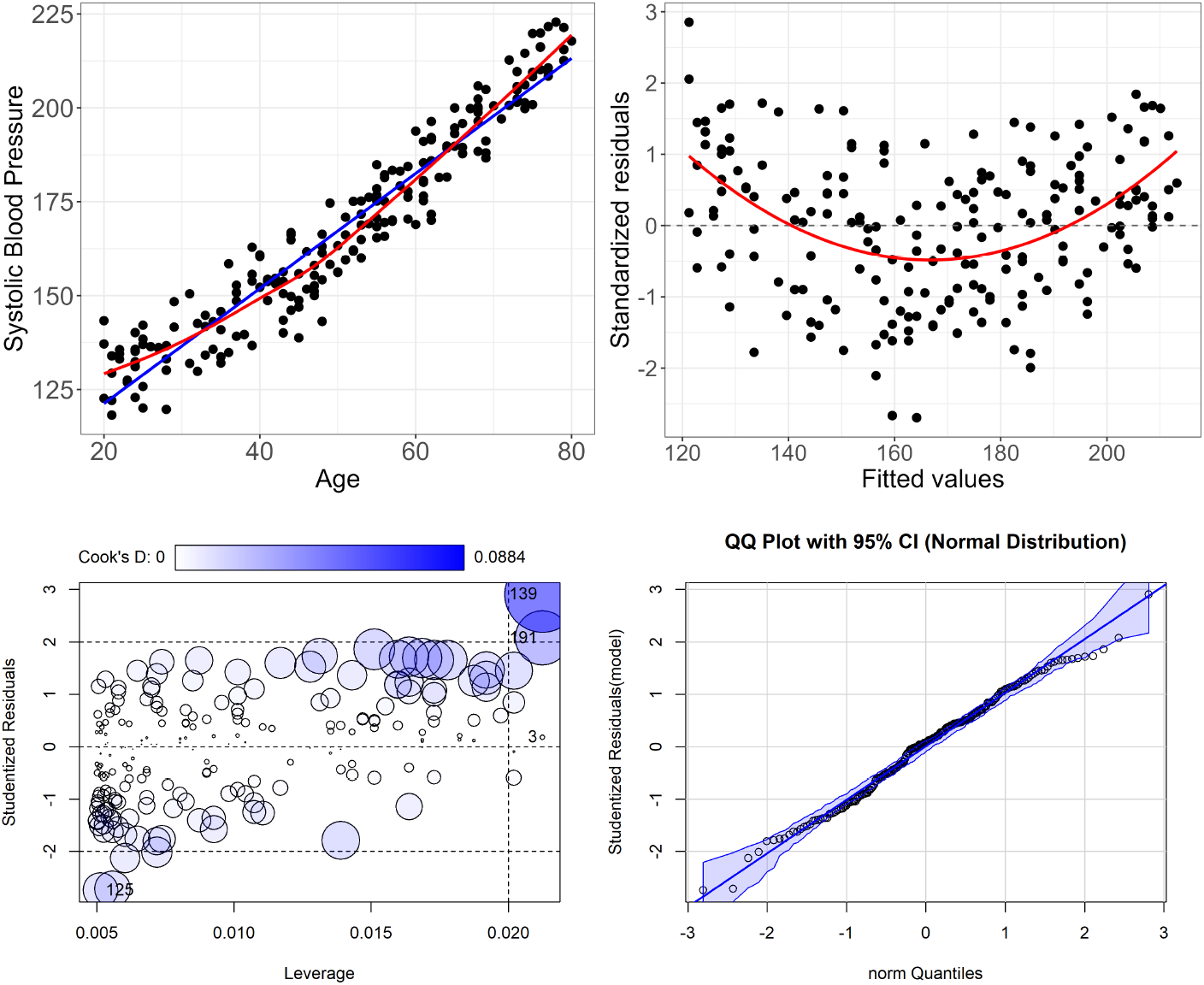
Simulated example showing a hypothetical relationship between blood pressure and age. The top-left scatter plot shows a clear positive relationship between age and blood pressure, with the blue line showing a linear fit and the red line a generalized additive model (GAM) fit. The residual plot (top-right) indicates a quadratic pattern that is not captured by the linear model but does not show evidence of heteroscedasticity. The Cook’s distance plot (bottom-left) shows that observations 139 and 191 exert greater influence on the model, though the overall Cook’s distances remain low, suggesting that outliers are not a major concern. The Q–Q plot (bottom-right) indicates that the residuals from the linear fit are approximately normally distributed.

Visualisations of the model were generated using the ‘visreg’ package in R [21]. For linear predictors, each plot displays the relationship between the predictor (x-axis) and the outcome (y-axis), with other covariates held constant. Results for each paper are available on GitHub [20]. Each regression analysis was assessed using the steps outlined below:

### Is the linear structure of the regression model appropriately specified?

- Scatterplots of standardized residuals versus predicted values and versus each predictor were used to assess the linearity of the overall model. Quadratic fits were overlaid to detect potential non-linear patterns [22].
- Residual specification tests for predictors were conducted using quadratic terms. For the overall fitted values, Tukey’s one-degree-of-freedom test was used to assess curvature and non-additivity [22].
- Scatterplots of raw dependent and predictor variables were examined with overlaid linear fits and smoothed generalized additive model (GAM) fits to explore potential non-linear relationships [23].

### Do residuals display homogeneity of variance?

- The plot of residuals vs predicted values was assessed for patterns such as funnelling.
- Plots of residuals vs individual variables were used to assess patterns.
- Heteroscedasticity was assessed using the studentized Breusch–Pagan test for non constant error variance [24].

### Were there outliers and influential observations?

- Cook’s distance measured the overall change in fit if the *ith* observation is removed. Potential influential observations are identified by Cook’s Distance*_i_ >* <u>^4^</u> , where *n* is the number of observations [25]. A threshold of 0.5 was used to identify influential observations.
- Difference in fitted values (DFFITS) were used to assess how many standard deviations the fitted value for an observation changes when that observation is removed from the model [25]. A practical cut-off of 1 was used to identify observations with substantial influence.
- Differences in beta estimates (DFBETAS) were used to assess how many standard errors a regression coefficient changes when an observation is removed from the model [25]. A practical cut-off of 1 was used to flag observations with a meaningful impact.
- Influence plots were examined to identify observations with both high leverage and large studentized residuals (typically ±2 or ±3), as these may disproportionately affect the model’s estimates. Such points appear as large, dark blue bubbles in the plots [22].
- The covariance ratio (COVRATIO) measures the overall change in the covariance matrix of the estimated regression coefficients when each observation is removed, reflecting changes in the precision of the coefficient estimates [26]. Values close to 1 indicate little influence on the model’s precision. Values below 1 indicate inflated variances and reduced precision (wider confidence intervals), whereas values above 1 indicate deflated variances and increased precision (narrower confidence intervals). A practical cut-off between 0.9 and 1.1 was applied, with values outside this range were considered to have a meaningful impact on precision.

### Are the residuals of the regression model approximately normally distributed?

- A Q–Q plot of model residuals with confidence bands assessed how residuals compared to the normal distribution. Bands indicated expected variability, and points outside the bands suggested systematic deviations [22].
- Residuals were described using descriptive statistics, including mean, standard deviation, median, minimum, maximum, skewness, and kurtosis.
- Normality was assessed using the Shapiro–Wilk and Kolmogorov–Smirnov tests [27], alongside graphical and descriptive diagnostics. Recognising that such tests are highly sensitive in large samples and may detect minor deviations, while lacking power in small samples, statistical significance alone was not taken as evidence of a meaningful violation. While formal tests contributed to the overall assessment, they were also examined to understand how often they reject minor departures from normality.

### Are the residuals of the model independent of each other?

- Each paper’s study design was reviewed to assess whether independence was theoretically plausible, with particular attention to potential clustering e.g., repeated measures within individuals or observations nested within hospitals.
- Serial correlation was assessed using the Durbin–Watson test [28].

### Is there collinearity among the independent variables?

- Variance Inflation Factors (VIFs) were examined to assess multicollinearity [28], with values greater than five considered potentially problematic [29].
- Changes or inflation in standard errors across regression models were monitored as indicators of multicollinearity or model instability.

### Inferential Reproducibility

Inferential reproducibility was assessed only for models that demonstrated sufficient computational agreement with the original results, defined as numerical agreement within pre-specified tolerance thresholds based on reported decimal precision [16]. Models classified as “Reproduced”, “Mostly reproduced”, or “Partially reproduced” were included, whereas models classified as “Not reproduced” were excluded as evaluating inferential reproducibility is of limited value when there were concerns about the data and/or methods. Models categorised as Partially reproduced were retained because they generally reflected the core structure of the original model, allowing assessment of whether violations of assumptions could plausibly have influenced the study conclusions. Inferential reproducibility was assessed as a binary (Yes/No) classification indicating whether the original inference remained credible after an assumption violation had been addressed.

Results were presented in HTML reports with four tabs: *Original Results*, *Reproduced Results*, *Differences*, and *Sensitivity Analysis*, allowing readers to navigate easily and compare findings. The reports are publicly available at https://github.com/Lee-V-Jones/Reproducibility/blob/main/README.md [20]. *Original results* were extracted from the text and tables of the published articles, with any unreported statistics left blank to highlight gaps. The *Reproduced Results* tab displayed re-estimated coefficients and model fit statistics (e.g., R^2^), along with a summary of model assumption checks. The *Differences* tab examined the agreement between original and reproduced results (e.g., regression coefficients, standard errors, confidence intervals, test statistics) which were assessed using tolerance thresholds based on reported decimal place precision, classifying values as “Reproduced”, “Incorrect rounding”, or “Not reproduced”. For computational reproducibility results, see Jones et al. [20].

The *Sensitivity Analysis* tab of the HTML report presents sensitivity analyses used to assess inferential reproducibility. Continuous dependent and independent variables from the reproduced models were standardized by centering and scaling to enable scale-free comparison of effect sizes. Standardized coefficients represent the expected change in the outcome, expressed in standard deviation units, for a one standard deviation increase in the predictor. Categorical variables were not standardized and were left in their original form (dummy variables). Standardization allowed coefficients to be compared on the same scale; however, comparing coefficients was not always an appropriate indicator of non-reproducibility, particularly when key model assumptions, such as linearity or distributional assumptions, were violated.

Differences in standardised regression coefficients were evaluated by comparing the reproduced model with sensitivity analyses assessing model assumptions. Three thresholds were applied: a percentage change of 10%, and absolute differences of 0.1 (moderate) and 0.2 (substantial). The 10% threshold was included as it was specified in the study protocol [30] and has been used in several previous studies [31, 32]. However, the percentage change can be misleading when regression coefficients are close to zero, as small absolute differences can appear disproportionately large. Because this was an exploratory project, we expanded on our protocol by also including absolute differences of 0.1 and 0.2 as more meaningful standardized measures. A difference of 0.1 can be considered similar in magnitude to a 10% change for many coefficients, while 0.2 aligns conceptually with Cohen’s d, where values below 0.2 are typically regarded as small [33]. Both percentage and absolute comparisons were explored to determine which thresholds more effectively captured meaningful differences and potential violations of assumptions.

There were five general steps followed to assess inferential reproducibility:

1. **Independence design/cluster effects**: If analyses were suspected of violating independence and an identification or clustering variable was available, the reproduced linear model (LM) and linear mixed model (LMM) were fit to compare the fixed effects [34]. The intraclass correlation coefficient (ICC) from the null LMM was reported [35]. Maximum-likelihood Akaike Information Criterion (AIC) values were then compared; when the LMM had a lower or similar AIC, it was retained as the primary model to preserve the data’s correlation structure for coefficient comparisons.
2. **Non-linearity**: Linearity was assessed by adding quadratic terms (or other non-linear terms as required). Non-linear terms were retained only when they substantially improved model fit, as indicated by reductions in AIC and Bayesian Information Criterion (BIC) values and by visual inspection that showed a better representation of the observed relationship.
3. **Bootstrap comparisons:** Confidence intervals (95% CIs) for linear models were estimated using the bias-corrected and accelerated (BCa) bootstrap method [36]. Where BCa intervals failed to converge, percentile bootstrap intervals were used. For outcomes with non-independent observations, linear mixed models (LMMs) were fitted [34], and CIs were obtained using a parametric bootstrap based on model simulations (10,000 resamples). When heteroscedasticity was suspected, a wild bootstrap was applied in both LM and LMM settings. Bootstrap distributions were inspected for bimodality and heavy tails; where results appeared sensitive to outliers, additional analyses were conducted to assess the influence of those observations on the estimated effects and resulting inferences.
4. **Comparison of results across models**: Percentage and absolute changes in estimates and confidence-interval bounds relative to the reproduced linear model were summarised using thresholds of 10% change and standardized coefficient differences of <0.10 and <0.20. Coefficient direction and statistical significance were assessed for consistency.
5. **Distribution check**: If inspection of model residuals or mean–variance relationships suggested that the assumed error distribution was inappropriate, sensitivity analyses were conducted using alternative distributions (e.g., Gamma for positively skewed continuous outcomes, Poisson or Negative Binomial for count data), as appropriate. Model assumptions, including the influence of outliers, were reassessed following refitting [37].

To address assumption violations, we prioritised simple, consistent approaches that enabled meaningful comparisons across models. Bootstrapping was the primary method used to account for violations of the normality assumption, with wild bootstrap [38] applied when heteroscedasticity was suspected. When linearity was questionable, models were refitted with polynomial terms, and model fit statistics such as R^2^ and AIC/BIC, were compared rather than coefficient estimates. In cases where residuals were non-normal or showed a clear mean–variance relationship, models based on alternative theoretical distributions (e.g., Poisson, Gamma) were fitted [23]. Model fit was compared using the AIC and BIC. When multicollinearity was detected, regularisation methods such as ridge regression were applied to obtain more stable and reliable coefficient estimates [39].

When studies reported removing outliers, we did not re-include them. This decision was based on two considerations: documentation surrounding outlier removal was often poor, and data for the outlier may not have been included in the dataset. To ensure valid comparisons of model fit, models were reproduced using the same dataset used for the original analysis. Recommendations regarding outlier handling were noted when relevant. Where necessary, alternative models were compared using information criteria, interpreted according to standard guidelines. For AIC, a reduction of less than 2 indicates support of a similar model, a reduction of 4–7 indicates improved fit, and a reduction greater than 10 indicates substantially better fit [40]. For BIC, Δ BIC values of 0–2 indicate weak evidence, 2–6 positive evidence, 6–10 strong evidence, and values greater than 10 very strong evidence in favour of the model with the lower BIC [41]. These thresholds are intended as general guidelines and were interpreted in the context of model purpose, plausibility, and diagnostic assessment.

Due to our limited contextual knowledge of the original studies and our decision not to contact the authors, we retained the variables reported in each model, even when the models appeared to be overfitted. In such cases, a recommendation to reduce the number of variables was noted. Models were classified as not inferentially reproducible when insufficient detail or key variables related to the study design were omitted, such as variables involving clustering, randomisation, or interactions. While efforts were made to standardize the overall approach, statistical analysis is inherently context-dependent. No single method is appropriate in all situations, and care was taken to select the most suitable approach for each case.

### Statistical Methods

The purpose of this study was descriptive rather than hypothesis test-driven. Descriptive statistics, including frequencies and percentages, were reported. Model-level classification of inferential reproducibility (Yes/No) was based on defined decision criteria. A standardized difference of 0.1 was used as the primary quantitative threshold, while a 10% change in key estimates and a standardized difference of 0.2 were reported descriptively for comparison. These quantitative criteria were considered alongside assessment of model assumptions (e.g., linearity and independence); the full decision framework is described above in the Inferential reproducibility section. A visualisation of the inferential reproducibility process is in Fig 2.

**Figure 2.**
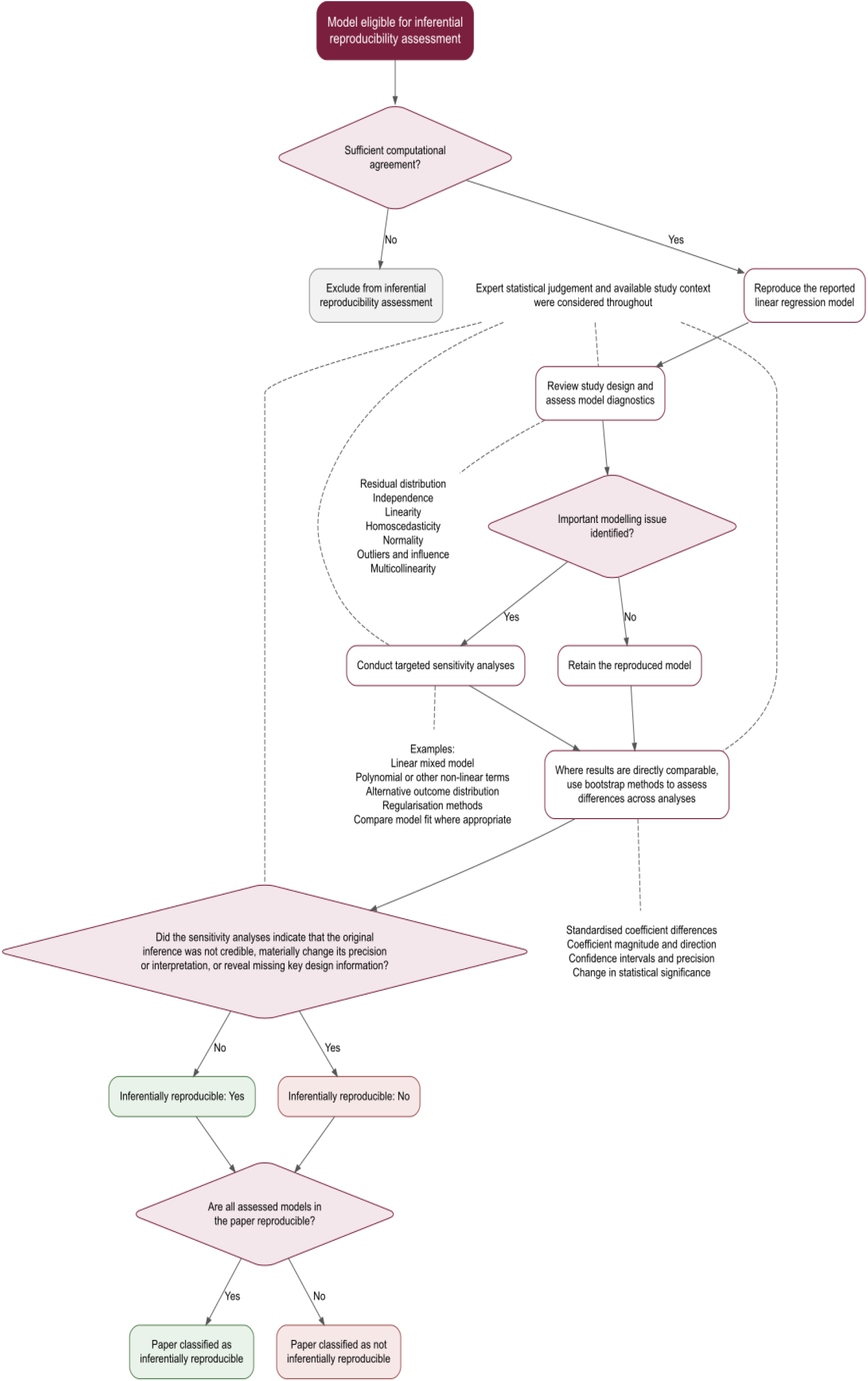
Flowchart for assessing inferential reproducibility. At the model level, up to three models were assessed for inferential reproducibility per paper, resulting in clustering of models within papers so independence could not be assumed. Accordingly, a Bayesian mixed-effects logistic regression model for the binary outcome of inferential reproducibility was fitted, with a random intercept for paper, to estimate predicted probabilities. Weakly informative priors were used to stabilise estimation given the small number of papers [42]. The intraclass correlation coefficient (ICC) and its 95% credible interval were derived from the posterior distribution of the paper-level random effect using the standard latent-scale formulation for logistic mixed models [43]. Model adequacy and convergence were assessed using standard Bayesian diagnostics, including rank-normalised convergence statistics and trace plots [44]. At the paper level, inferential reproducibility was summarised as the proportion of papers in which all assessed models met the reproducibility criterion, with 95% Wilson confidence intervals used to quantify uncertainty around this estimate. R version 4.4.2 was used for all analyses [27].

## Results

Of the 95 papers reviewed, 80 (84%) were observational studies, and 73 (77%) involved human participants. Among these papers, 68 reported having data available; however, the raw data necessary to reproduce the analyses could not be identified in 25 papers [16]. From the original random sample, a subset of 20 papers were sequentially assessed for computational reproducibility, of which eight were successfully reproduced, with a further six papers containing models that were at least partially reproducible. Therefore, 14 papers were evaluated for inferential reproducibility. Of these, three or 21% (95% CI: 8%, 48%) met the inferential reproducibility criterion, defined as all assessed models within the paper being reproducible (Fig 3).

**Figure 3.**
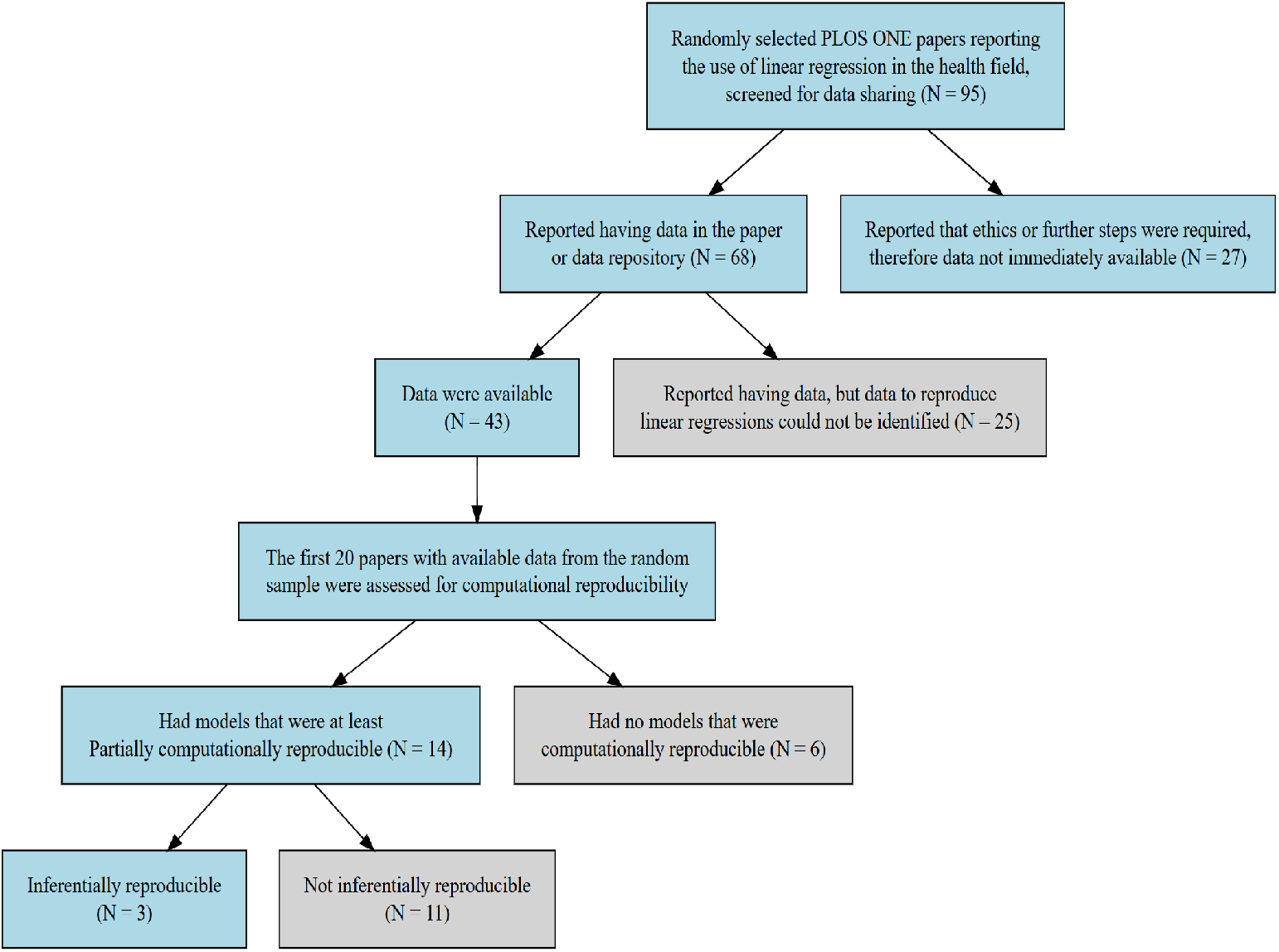
Flowchart of the included papers in the analysis. N is the number of papers.

In total, 32 models across 14 papers were assessed for inferential reproducibility. Of these models, 14/32 (44%) were inferentially reproducible based on the observed data. After accounting for clustering by paper, the model-based marginal probability of reproducibility was 0.34 (95% Credible Intervals 0.07, 0.69). The reduction relative to the observed proportion reflects substantial between-paper variation, with inferential reproducibility tending to cluster within papers (ICC = 0.59, 95% CI: 0.05, 0.89). Given the small number of papers (N = 14) and the resulting uncertainty reflected in the wide credible interval, the estimate should be interpreted cautiously. Diagnostics for the Bayesian model used in this study indicated satisfactory convergence, and posterior predictive checks suggested adequate representation of the observed data. Full model results and diagnostic plots are provided in [20].

Inferential reproducibility was assessed primarily by examining whether regression coefficients and their 95% confidence intervals fell within specified thresholds. This approach recognises that departures from model assumptions may arise from multiple interacting sources; therefore, specific violations cannot always be directly linked to discrepancies in estimates unless the connection is clear. Models with minor assumption departures were still considered inferentially reproducible when diagnostic assessments indicated these were unlikely to materially affect parameter estimates, interpretation, or overall model adequacy.

Most authors reported limited information on statistical assumptions. Table 1 summarises the number of papers in which assumptions were checked or discussed, and the number in which at least one violation was identified in the reproduced analyses. Sample sizes differ between the reported and reproduced analyses: the reported results reflect the full paper, whereas the reproduced analyses were restricted to the three selected models per paper.

**Table 1.** Statistical assumption checks reported by authors and our assessment of violations across papers.

| Assumptions | Paper reported |  | Reproduced analyses |  |
| --- | --- | --- | --- | --- |
|  | N | Checked | N | Violated |
| Normality | 14 | 8 (57%) | 14 | 8 (57%) |
| Linearity | 12 | 2 (17%) | 11 | 5 (45%) |
| Homoscedasticity | 14 | 3 (21%) | 13 | 7 (54%) |
| Independence | 14 | 2 (14%) | 14 | 8 (57%) |
| Collinearity | 11 | 4 (36%) | 8 | 0 (0%) |
| Outlier | 14 | 6 (43%) | 14 | 4 (29%) |

Paper reported: Authors reported checking or discussing the assumption; Reproduced analyses: Assumption violated in at least one of our reproduced analyses.

Residual normality was assessed using the Kolmogorov–Smirnov (KS) and Shapiro–Wilk (SW) tests, with Q-Q plots informing the final assessment across 32 models. In five models, all tests and plots indicated there was no violation of normality; in seven models, all indicated there were violations of normality. In seven models, one of the tests indicated a violation and the Q–Q plot also suggested non-normality. There was 13 cases where one of the tests indicated a violation, but the deviation on the Q-Q plot was negligible, and the residuals were judged to be approximately normal.

The most frequently violated assumptions were residual normality and independence, each of which was violated in eight papers. Mild departures from normality did not always lead to inferential failure; however, violations of independence almost always did, with 16/32 models exhibiting independence violations. For descriptions and reproducibility results for individual papers, see Table 2.

**Table 2.**
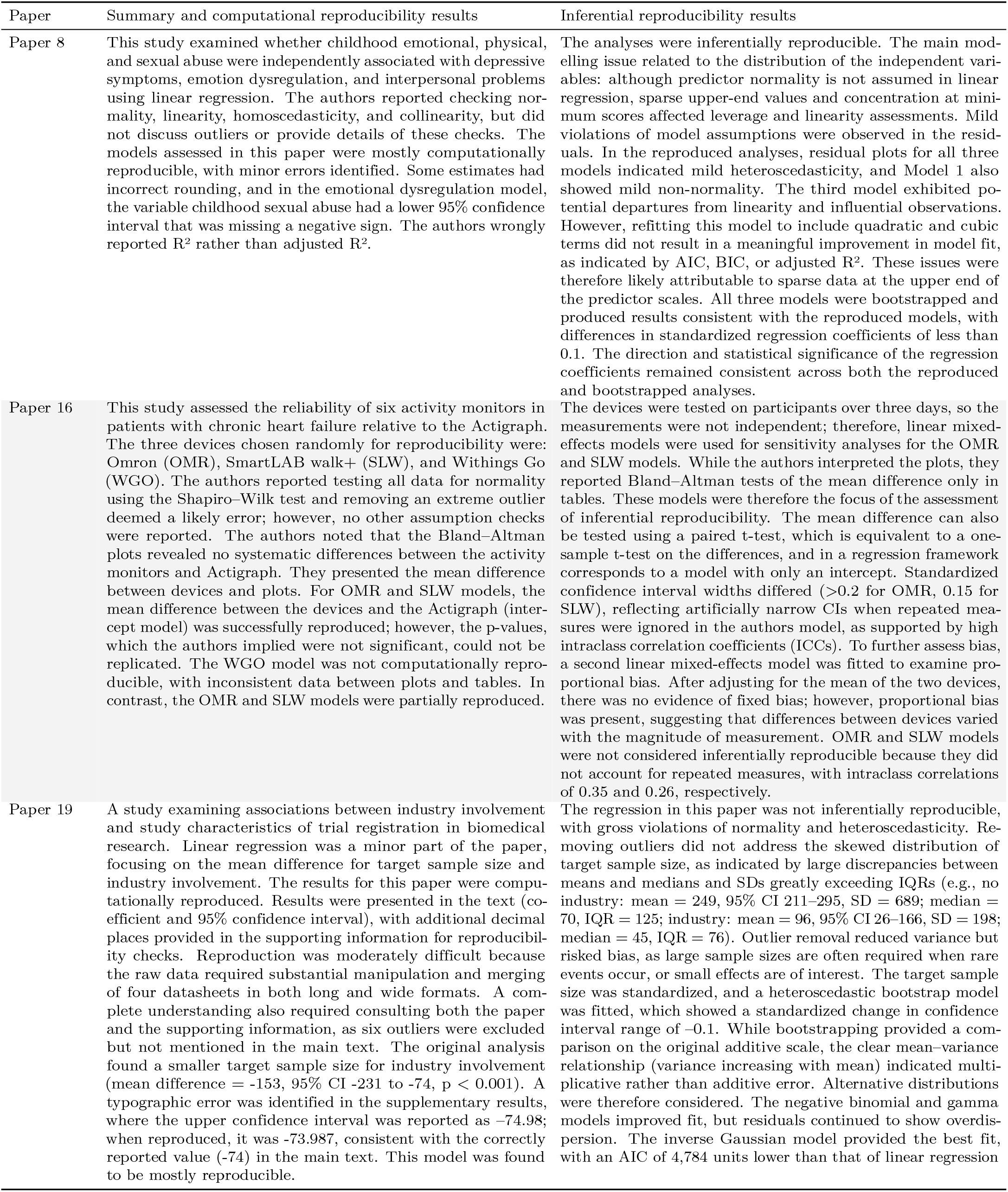
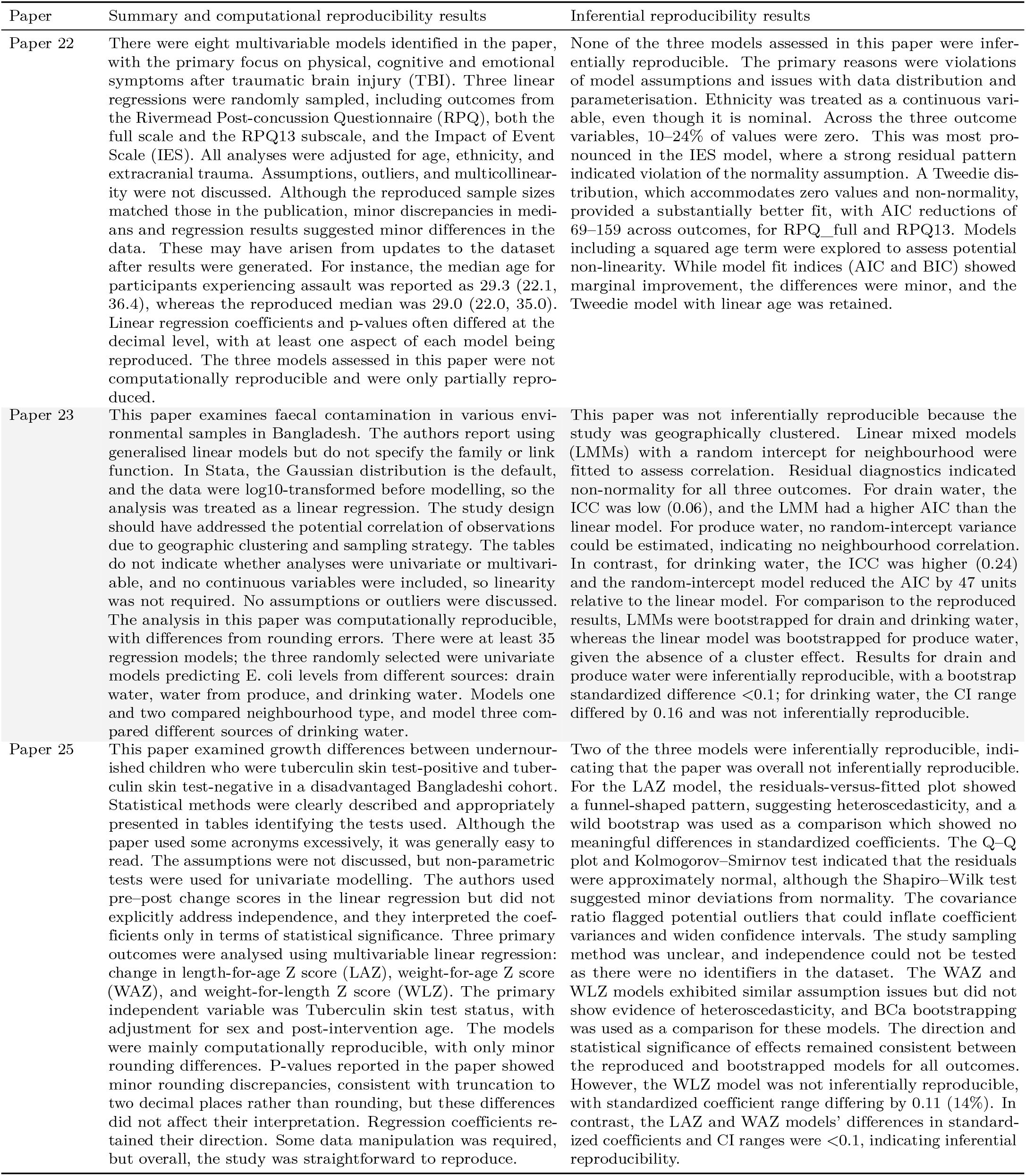
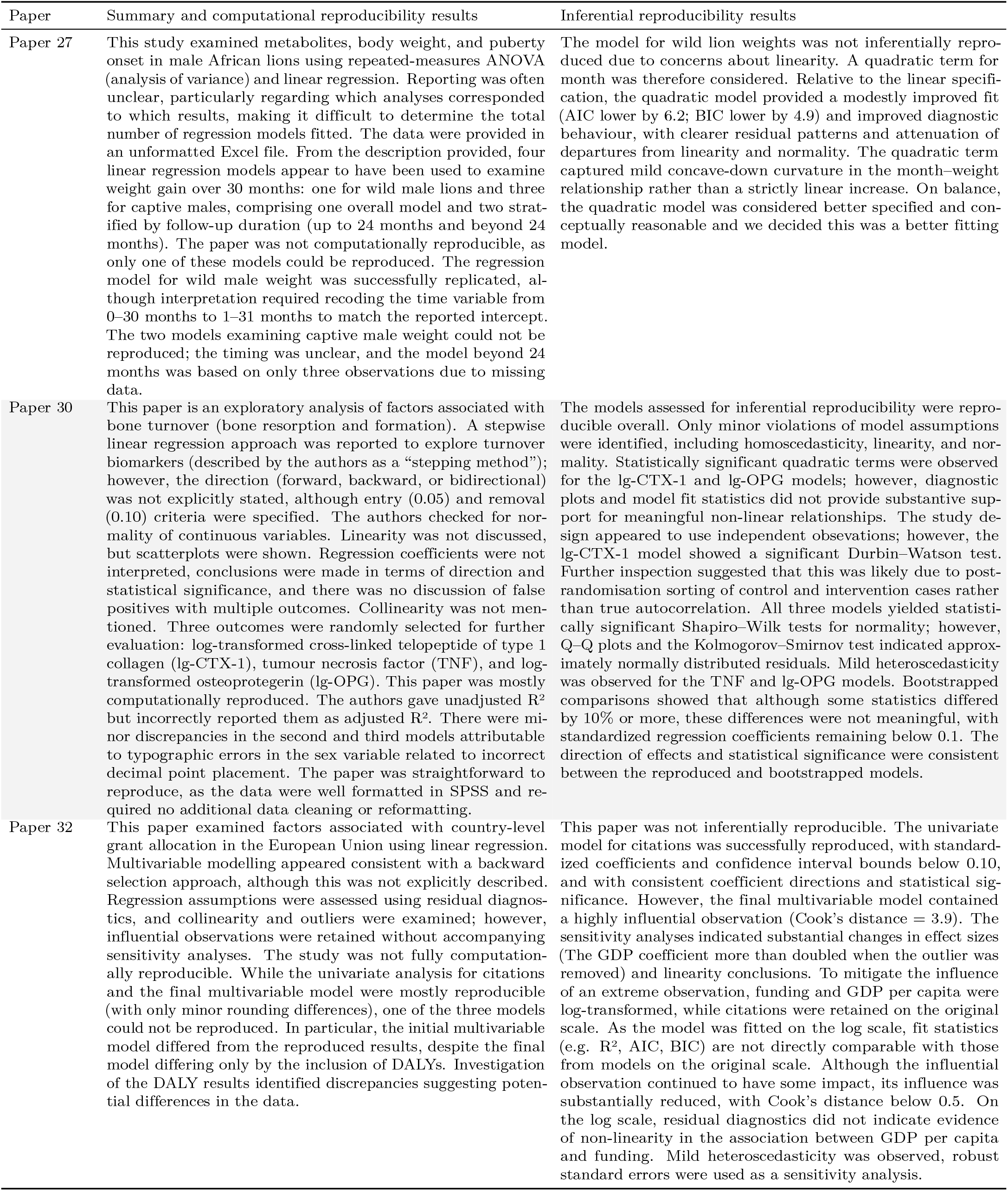
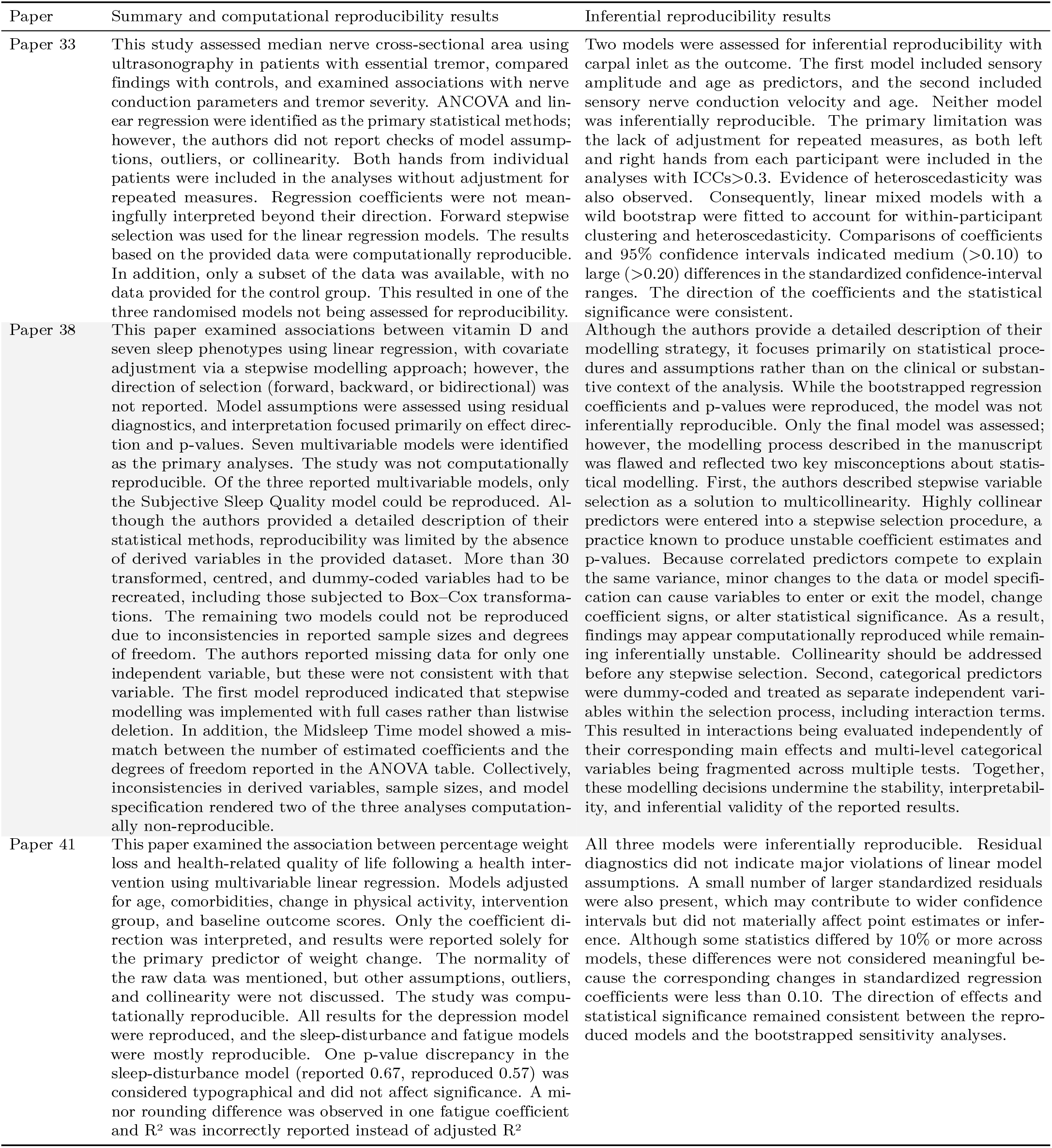
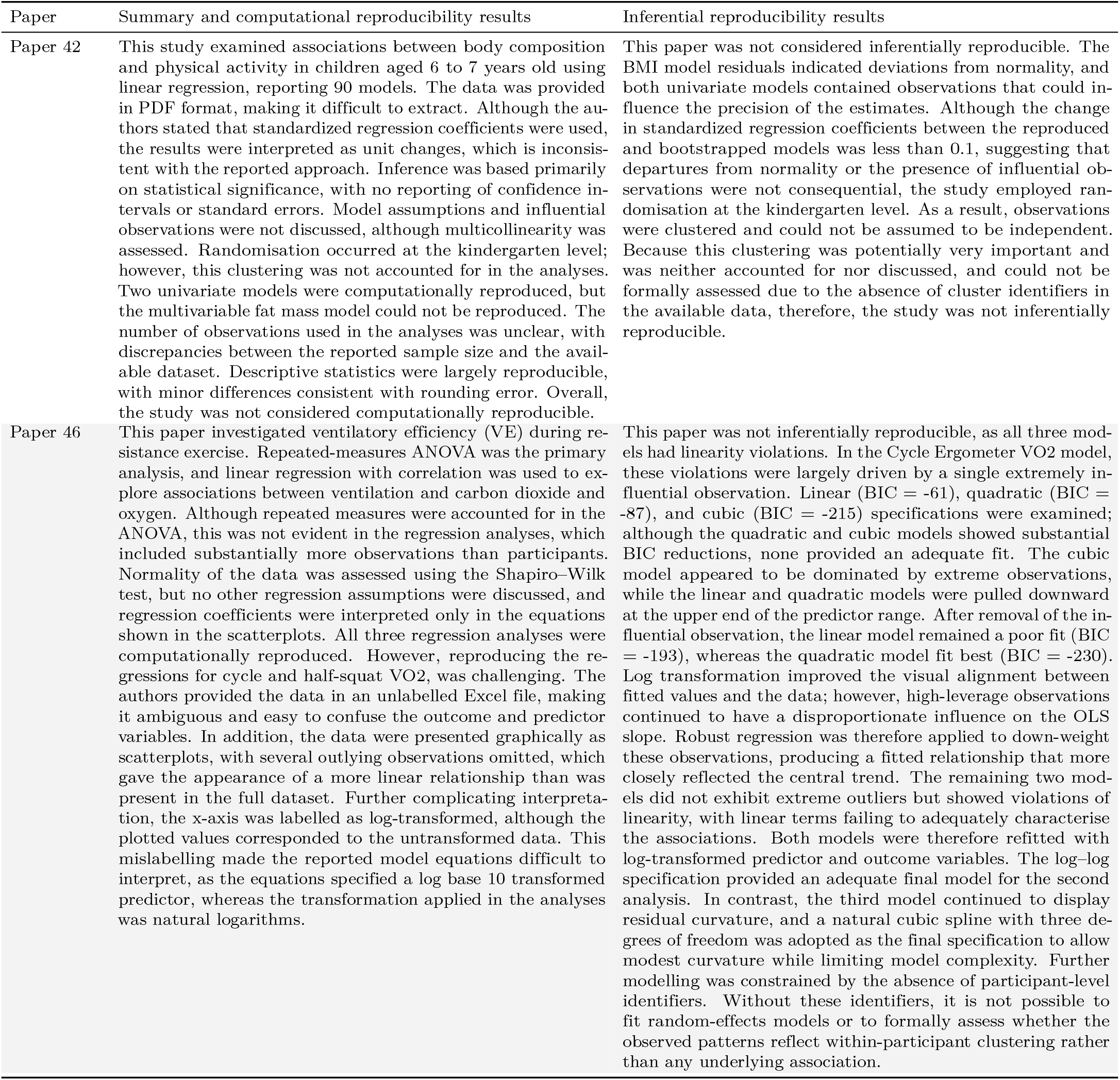
Summary and paper-level reproducibility results.

| Paper | Summary and computational reproducibility results | Inferential reproducibility results |
| --- | --- | --- |
| Paper 8 | This study examined whether childhood emotional, physical, and sexual abuse were independently associated with depressive symptoms, emotion dysregulation, and interpersonal problems using linear regression. The authors reported checking normality, linearity, homoscedasticity, and collinearity, but did not discuss outliers or provide details of these checks. The models assessed in this paper were mostly computationally reproducible, with minor errors identified. Some estimates had incorrect rounding, and in the emotional dysregulation model, the variable childhood sexual abuse had a lower 95% confidence interval that was missing a negative sign. The authors wrongly reported $R^2$ rather than adjusted $R^2$. | The analyses were inferentially reproducible. The main modelling issue related to the distribution of the independent variables: although predictor normality is not assumed in linear regression, sparse upper-end values and concentration at minimum scores affected leverage and linearity assessments. Mild violations of model assumptions were observed in the residuals. In the reproduced analyses, residual plots for all three models indicated mild heteroscedasticity, and Model 1 also showed mild non-normality. The third model exhibited potential departures from linearity and influential observations. However, refitting this model to include quadratic and cubic terms did not result in a meaningful improvement in model fit, as indicated by AIC, BIC, or adjusted $R^2$. These issues were therefore likely attributable to sparse data at the upper end of the predictor scales. All three models were bootstrapped and produced results consistent with the reproduced models, with differences in standardized regression coefficients of less than 0.1. The direction and statistical significance of the regression coefficients remained consistent across both the reproduced and bootstrapped analyses. |
| Paper 16 | This study assessed the reliability of six activity monitors in patients with chronic heart failure relative to the Actigraph. The three devices chosen randomly for reproducibility were: Omron (OMR), SmartLAB walk+ (SLW), and Withings Go (WGO). The authors reported testing all data for normality using the Shapiro–Wilk test and removing an extreme outlier deemed a likely error; however, no other assumption checks were reported. The authors noted that the Bland–Altman plots revealed no systematic differences between the activity monitors and Actigraph. They presented the mean difference between devices and plots. For OMR and SLW models, the mean difference between the devices and the Actigraph (intercept model) was successfully reproduced; however, the p-values, which the authors implied were not significant, could not be replicated. The WGO model was not computationally reproducible, with inconsistent data between plots and tables. In contrast, the OMR and SLW models were partially reproduced. | The devices were tested on participants over three days, so the measurements were not independent; therefore, linear mixed-effects models were used for sensitivity analyses for the OMR and SLW models. While the authors interpreted the plots, they reported Bland–Altman tests of the mean difference only in tables. These models were therefore the focus of the assessment of inferential reproducibility. The mean difference can also be tested using a paired t-test, which is equivalent to a one-sample t-test on the differences, and in a regression framework corresponds to a model with only an intercept. Standardized confidence interval widths differed ($>0.2$ for OMR, $0.15$ for SLW), reflecting artificially narrow CIs when repeated measures were ignored in the authors model, as supported by high intraclass correlation coefficients (ICCs). To further assess bias, a second linear mixed-effects model was fitted to examine proportional bias. After adjusting for the mean of the two devices, there was no evidence of fixed bias; however, proportional bias was present, suggesting that differences between devices varied with the magnitude of measurement. OMR and SLW models were not considered inferentially reproducible because they did not account for repeated measures, with intraclass correlations of $0.35$ and $0.26$, respectively. |
| Paper 19 | A study examining associations between industry involvement and study characteristics of trial registration in biomedical research. Linear regression was a minor part of the paper, focusing on the mean difference for target sample size and industry involvement. The results for this paper were computationally reproduced. Results were presented in the text (coefficient and 95% confidence interval), with additional decimal places provided in the supporting information for reproducibility checks. Reproduction was moderately difficult because the raw data required substantial manipulation and merging of four datasheets in both long and wide formats. A complete understanding also required consulting both the paper and the supporting information, as six outliers were excluded but not mentioned in the main text. The original analysis found a smaller target sample size for industry involvement (mean difference = $-153$, 95% CI $-231$ to $-74$, $p < 0.001$). A typographic error was identified in the supplementary results, where the upper confidence interval was reported as $-74.98$; when reproduced, it was $-73.987$, consistent with the correctly reported value ($-74$) in the main text. This model was found to be mostly reproducible. | The regression in this paper was not inferentially reproducible, with gross violations of normality and heteroscedasticity. Removing outliers did not address the skewed distribution of target sample size, as indicated by large discrepancies between means and medians and SDs greatly exceeding IQRs (e.g., no industry: mean = $249$, 95% CI $211$–$295$, SD = $689$; median = $70$, IQR = $125$; industry: mean = $96$, 95% CI $26$–$166$, SD = $198$; median = $45$, IQR = $76$). Outlier removal reduced variance but risked bias, as large sample sizes are often required when rare events occur, or small effects are of interest. The target sample size was standardized, and a heteroscedastic bootstrap model was fitted, which showed a standardized change in confidence interval range of $-0.1$. While bootstrapping provided a comparison on the original additive scale, the clear mean–variance relationship (variance increasing with mean) indicated multiplicative rather than additive error. Alternative distributions were therefore considered. The negative binomial and gamma models improved fit, but residuals continued to show overdispersion. The inverse Gaussian model provided the best fit, with an AIC of $4,784$ units lower than that of linear regression |
| Paper 22 | There were eight multivariable models identified in the paper, with the primary focus on physical, cognitive and emotional symptoms after traumatic brain injury (TBI). Three linear regressions were randomly sampled, including outcomes from the Rivermead Post-concussion Questionnaire (RPQ), both the full scale and the RPQ13 subscale, and the Impact of Event Scale (IES). All analyses were adjusted for age, ethnicity, and extracranial trauma. Assumptions, outliers, and multicollinearity were not discussed. Although the reproduced sample sizes matched those in the publication, minor discrepancies in medians and regression results suggested minor differences in the data. These may have arisen from updates to the dataset after results were generated. For instance, the median age for participants experiencing assault was reported as 29.3 (22.1, 36.4), whereas the reproduced median was 29.0 (22.0, 35.0). Linear regression coefficients and p-values often differed at the decimal level, with at least one aspect of each model being reproduced. The three models assessed in this paper were not computationally reproducible and were only partially reproduced. | None of the three models assessed in this paper were inferentially reproducible. The primary reasons were violations of model assumptions and issues with data distribution and parameterisation. Ethnicity was treated as a continuous variable, even though it is nominal. Across the three outcome variables, 10–24% of values were zero. This was most pronounced in the IES model, where a strong residual pattern indicated violation of the normality assumption. A Tweedie distribution, which accommodates zero values and non-normality, provided a substantially better fit, with AIC reductions of 69–159 across outcomes, for RPQ_full and RPQ13. Models including a squared age term were explored to assess potential non-linearity. While model fit indices (AIC and BIC) showed marginal improvement, the differences were minor, and the Tweedie model with linear age was retained. |
| Paper 23 | This paper examines faecal contamination in various environmental samples in Bangladesh. The authors report using generalised linear models but do not specify the family or link function. In Stata, the Gaussian distribution is the default, and the data were log10-transformed before modelling, so the analysis was treated as a linear regression. The study design should have addressed the potential correlation of observations due to geographic clustering and sampling strategy. The tables do not indicate whether analyses were univariate or multivariable, and no continuous variables were included, so linearity was not required. No assumptions or outliers were discussed. The analysis in this paper was computationally reproducible, with differences from rounding errors. There were at least 35 regression models; the three randomly selected were univariate models predicting *E. coli* levels from different sources: drain water, water from produce, and drinking water. Models one and two compared neighbourhood type, and model three compared different sources of drinking water. | This paper was not inferentially reproducible because the study was geographically clustered. Linear mixed models (LMMs) with a random intercept for neighbourhood were fitted to assess correlation. Residual diagnostics indicated non-normality for all three outcomes. For drain water, the ICC was low (0.06), and the LMM had a higher AIC than the linear model. For produce water, no random-intercept variance could be estimated, indicating no neighbourhood correlation. In contrast, for drinking water, the ICC was higher (0.24) and the random-intercept model reduced the AIC by 47 units relative to the linear model. For comparison to the reproduced results, LMMs were bootstrapped for drain and drinking water, whereas the linear model was bootstrapped for produce water, given the absence of a cluster effect. Results for drain and produce water were inferentially reproducible, with a bootstrap standardized difference <0.1; for drinking water, the CI range differed by 0.16 and was not inferentially reproducible. |
| Paper 25 | This paper examined growth differences between undernourished children who were tuberculin skin test-positive and tuberculin skin test-negative in a disadvantaged Bangladeshi cohort. Statistical methods were clearly described and appropriately presented in tables identifying the tests used. Although the paper used some acronyms excessively, it was generally easy to read. The assumptions were not discussed, but non-parametric tests were used for univariate modelling. The authors used pre-post change scores in the linear regression but did not explicitly address independence, and they interpreted the coefficients only in terms of statistical significance. Three primary outcomes were analysed using multivariable linear regression: change in length-for-age Z score (LAZ), weight-for-age Z score (WAZ), and weight-for-length Z score (WLZ). The primary independent variable was Tuberculin skin test status, with adjustment for sex and post-intervention age. The models were mainly computationally reproducible, with only minor rounding differences. P-values reported in the paper showed minor rounding discrepancies, consistent with truncation to two decimal places rather than rounding, but these differences did not affect their interpretation. Regression coefficients retained their direction. Some data manipulation was required, but overall, the study was straightforward to reproduce. | Two of the three models were inferentially reproducible, indicating that the paper was overall not inferentially reproducible. For the LAZ model, the residuals-versus-fitted plot showed a funnel-shaped pattern, suggesting heteroscedasticity, and a wild bootstrap was used as a comparison which showed no meaningful differences in standardized coefficients. The Q-Q plot and Kolmogorov-Smirnov test indicated that the residuals were approximately normal, although the Shapiro-Wilk test suggested minor deviations from normality. The covariance ratio flagged potential outliers that could inflate coefficient variances and widen confidence intervals. The study sampling method was unclear, and independence could not be tested as there were no identifiers in the dataset. The WAZ and WLZ models exhibited similar assumption issues but did not show evidence of heteroscedasticity, and BCa bootstrapping was used as a comparison for these models. The direction and statistical significance of effects remained consistent between the reproduced and bootstrapped models for all outcomes. However, the WLZ model was not inferentially reproducible, with standardized coefficient range differing by 0.11 (14%). In contrast, the LAZ and WAZ models' differences in standardized coefficients and CI ranges were <0.1, indicating inferential reproducibility. |
| Paper 27 | This study examined metabolites, body weight, and puberty onset in male African lions using repeated-measures ANOVA (analysis of variance) and linear regression. Reporting was often unclear, particularly regarding which analyses corresponded to which results, making it difficult to determine the total number of regression models fitted. The data were provided in an unformatted Excel file. From the description provided, four linear regression models appear to have been used to examine weight gain over 30 months: one for wild male lions and three for captive males, comprising one overall model and two stratified by follow-up duration (up to 24 months and beyond 24 months). The paper was not computationally reproducible, as only one of these models could be reproduced. The regression model for wild male weight was successfully replicated, although interpretation required recoding the time variable from 0–30 months to 1–31 months to match the reported intercept. The two models examining captive male weight could not be reproduced; the timing was unclear, and the model beyond 24 months was based on only three observations due to missing data. | The model for wild lion weights was not inferentially reproduced due to concerns about linearity. A quadratic term for month was therefore considered. Relative to the linear specification, the quadratic model provided a modestly improved fit (AIC lower by 6.2; BIC lower by 4.9) and improved diagnostic behaviour, with clearer residual patterns and attenuation of departures from linearity and normality. The quadratic term captured mild concave-down curvature in the month–weight relationship rather than a strictly linear increase. On balance, the quadratic model was considered better specified and conceptually reasonable and we decided this was a better fitting model. |
| Paper 30 | This paper is an exploratory analysis of factors associated with bone turnover (bone resorption and formation). A stepwise linear regression approach was reported to explore turnover biomarkers (described by the authors as a “stepping method”); however, the direction (forward, backward, or bidirectional) was not explicitly stated, although entry (0.05) and removal (0.10) criteria were specified. The authors checked for normality of continuous variables. Linearity was not discussed, but scatterplots were shown. Regression coefficients were not interpreted, conclusions were made in terms of direction and statistical significance, and there was no discussion of false positives with multiple outcomes. Collinearity was not mentioned. Three outcomes were randomly selected for further evaluation: log-transformed cross-linked telopeptide of type 1 collagen (lg-CTX-1), tumour necrosis factor (TNF), and log-transformed osteoprotegerin (lg-OPG). This paper was mostly computationally reproduced. The authors gave unadjusted R<sup>2</sup> but incorrectly reported them as adjusted R<sup>2</sup>. There were minor discrepancies in the second and third models attributable to typographic errors in the sex variable related to incorrect decimal point placement. The paper was straightforward to reproduce, as the data were well formatted in SPSS and required no additional data cleaning or reformatting. | The models assessed for inferential reproducibility were reproducible overall. Only minor violations of model assumptions were identified, including homoscedasticity, linearity, and normality. Statistically significant quadratic terms were observed for the lg-CTX-1 and lg-OPG models; however, diagnostic plots and model fit statistics did not provide substantive support for meaningful non-linear relationships. The study design appeared to use independent observations; however, the lg-CTX-1 model showed a significant Durbin–Watson test. Further inspection suggested that this was likely due to post-randomisation sorting of control and intervention cases rather than true autocorrelation. All three models yielded statistically significant Shapiro–Wilk tests for normality; however, Q–Q plots and the Kolmogorov–Smirnov test indicated approximately normally distributed residuals. Mild heteroscedasticity was observed for the TNF and lg-OPG models. Bootstrapped comparisons showed that although some statistics differed by 10% or more, these differences were not meaningful, with standardized regression coefficients remaining below 0.1. The direction of effects and statistical significance were consistent between the reproduced and bootstrapped models. |
| Paper 32 | This paper examined factors associated with country-level grant allocation in the European Union using linear regression. Multivariable modelling appeared consistent with a backward selection approach, although this was not explicitly described. Regression assumptions were assessed using residual diagnostics, and collinearity and outliers were examined; however, influential observations were retained without accompanying sensitivity analyses. The study was not fully computationally reproducible. While the univariate analysis for citations and the final multivariable model were mostly reproducible (with only minor rounding differences), one of the three models could not be reproduced. In particular, the initial multivariable model differed from the reproduced results, despite the final model differing only by the inclusion of DALYs. Investigation of the DALY results identified discrepancies suggesting potential differences in the data. | This paper was not inferentially reproducible. The univariate model for citations was successfully reproduced, with standardized coefficients and confidence interval bounds below 0.10, and with consistent coefficient directions and statistical significance. However, the final multivariable model contained a highly influential observation (Cook’s distance = 3.9). The sensitivity analyses indicated substantial changes in effect sizes (The GDP coefficient more than doubled when the outlier was removed) and linearity conclusions. To mitigate the influence of an extreme observation, funding and GDP per capita were log-transformed, while citations were retained on the original scale. As the model was fitted on the log scale, fit statistics (e.g. R<sup>2</sup>, AIC, BIC) are not directly comparable with those from models on the original scale. Although the influential observation continued to have some impact, its influence was substantially reduced, with Cook’s distance below 0.5. On the log scale, residual diagnostics did not indicate evidence of non-linearity in the association between GDP per capita and funding. Mild heteroscedasticity was observed, robust standard errors were used as a sensitivity analysis. |
| Paper 33 | This study assessed median nerve cross-sectional area using ultrasonography in patients with essential tremor, compared findings with controls, and examined associations with nerve conduction parameters and tremor severity. ANCOVA and linear regression were identified as the primary statistical methods; however, the authors did not report checks of model assumptions, outliers, or collinearity. Both hands from individual patients were included in the analyses without adjustment for repeated measures. Regression coefficients were not meaningfully interpreted beyond their direction. Forward stepwise selection was used for the linear regression models. The results based on the provided data were computationally reproducible. In addition, only a subset of the data was available, with no data provided for the control group. This resulted in one of the three randomised models not being assessed for reproducibility. | Two models were assessed for inferential reproducibility with carpal inlet as the outcome. The first model included sensory amplitude and age as predictors, and the second included sensory nerve conduction velocity and age. Neither model was inferentially reproducible. The primary limitation was the lack of adjustment for repeated measures, as both left and right hands from each participant were included in the analyses with ICCs>0.3. Evidence of heteroscedasticity was also observed. Consequently, linear mixed models with a wild bootstrap were fitted to account for within-participant clustering and heteroscedasticity. Comparisons of coefficients and 95% confidence intervals indicated medium (>0.10) to large (>0.20) differences in the standardized confidence-interval ranges. The direction of the coefficients and the statistical significance were consistent. |
| Paper 38 | This paper examined associations between vitamin D and seven sleep phenotypes using linear regression, with covariate adjustment via a stepwise modelling approach; however, the direction of selection (forward, backward, or bidirectional) was not reported. Model assumptions were assessed using residual diagnostics, and interpretation focused primarily on effect direction and p-values. Seven multivariable models were identified as the primary analyses. The study was not computationally reproducible. Of the three reported multivariable models, only the Subjective Sleep Quality model could be reproduced. Although the authors provided a detailed description of their statistical methods, reproducibility was limited by the absence of derived variables in the provided dataset. More than 30 transformed, centred, and dummy-coded variables had to be recreated, including those subjected to Box-Cox transformations. The remaining two models could not be reproduced due to inconsistencies in reported sample sizes and degrees of freedom. The authors reported missing data for only one independent variable, but these were not consistent with that variable. The first model reproduced indicated that stepwise modelling was implemented with full cases rather than listwise deletion. In addition, the Midsleep Time model showed a mismatch between the number of estimated coefficients and the degrees of freedom reported in the ANOVA table. Collectively, inconsistencies in derived variables, sample sizes, and model specification rendered two of the three analyses computationally non-reproducible. | Although the authors provide a detailed description of their modelling strategy, it focuses primarily on statistical procedures and assumptions rather than on the clinical or substantive context of the analysis. While the bootstrapped regression coefficients and p-values were reproduced, the model was not inferentially reproducible. Only the final model was assessed; however, the modelling process described in the manuscript was flawed and reflected two key misconceptions about statistical modelling. First, the authors described stepwise variable selection as a solution to multicollinearity. Highly collinear predictors were entered into a stepwise selection procedure, a practice known to produce unstable coefficient estimates and p-values. Because correlated predictors compete to explain the same variance, minor changes to the data or model specification can cause variables to enter or exit the model, change coefficient signs, or alter statistical significance. As a result, findings may appear computationally reproduced while remaining inferentially unstable. Collinearity should be addressed before any stepwise selection. Second, categorical predictors were dummy-coded and treated as separate independent variables within the selection process, including interaction terms. This resulted in interactions being evaluated independently of their corresponding main effects and multi-level categorical variables being fragmented across multiple tests. Together, these modelling decisions undermine the stability, interpretability, and inferential validity of the reported results. |
| Paper 41 | This paper examined the association between percentage weight loss and health-related quality of life following a health intervention using multivariable linear regression. Models adjusted for age, comorbidities, change in physical activity, intervention group, and baseline outcome scores. Only the coefficient direction was interpreted, and results were reported solely for the primary predictor of weight change. The normality of the raw data was mentioned, but other assumptions, outliers, and collinearity were not discussed. The study was computationally reproducible. All results for the depression model were reproduced, and the sleep-disturbance and fatigue models were mostly reproducible. One p-value discrepancy in the sleep-disturbance model (reported 0.67, reproduced 0.57) was considered typographical and did not affect significance. A minor rounding difference was observed in one fatigue coefficient and R<sup>2</sup> was incorrectly reported instead of adjusted R<sup>2</sup> | All three models were inferentially reproducible. Residual diagnostics did not indicate major violations of linear model assumptions. A small number of larger standardized residuals were also present, which may contribute to wider confidence intervals but did not materially affect point estimates or inference. Although some statistics differed by 10% or more across models, these differences were not considered meaningful because the corresponding changes in standardized regression coefficients were less than 0.10. The direction of effects and statistical significance remained consistent between the reproduced models and the bootstrapped sensitivity analyses. |
| Paper 42 | This study examined associations between body composition and physical activity in children aged 6 to 7 years old using linear regression, reporting 90 models. The data was provided in PDF format, making it difficult to extract. Although the authors stated that standardized regression coefficients were used, the results were interpreted as unit changes, which is inconsistent with the reported approach. Inference was based primarily on statistical significance, with no reporting of confidence intervals or standard errors. Model assumptions and influential observations were not discussed, although multicollinearity was assessed. Randomisation occurred at the kindergarten level; however, this clustering was not accounted for in the analyses. Two univariate models were computationally reproduced, but the multivariable fat mass model could not be reproduced. The number of observations used in the analyses was unclear, with discrepancies between the reported sample size and the available dataset. Descriptive statistics were largely reproducible, with minor differences consistent with rounding error. Overall, the study was not considered computationally reproducible. | This paper was not considered inferentially reproducible. The BMI model residuals indicated deviations from normality, and both univariate models contained observations that could influence the precision of the estimates. Although the change in standardized regression coefficients between the reproduced and bootstrapped models was less than 0.1, suggesting that departures from normality or the presence of influential observations were not consequential, the study employed randomisation at the kindergarten level. As a result, observations were clustered and could not be assumed to be independent. Because this clustering was potentially very important and was neither accounted for nor discussed, and could not be formally assessed due to the absence of cluster identifiers in the available data, therefore, the study was not inferentially reproducible. |
| Paper 46 | This paper investigated ventilatory efficiency (VE) during resistance exercise. Repeated-measures ANOVA was the primary analysis, and linear regression with correlation was used to explore associations between ventilation and carbon dioxide and oxygen. Although repeated measures were accounted for in the ANOVA, this was not evident in the regression analyses, which included substantially more observations than participants. Normality of the data was assessed using the Shapiro-Wilk test, but no other regression assumptions were discussed, and regression coefficients were interpreted only in the equations shown in the scatterplots. All three regression analyses were computationally reproduced. However, reproducing the regressions for cycle and half-squat VO<sub>2</sub>, was challenging. The authors provided the data in an unlabelled Excel file, making it ambiguous and easy to confuse the outcome and predictor variables. In addition, the data were presented graphically as scatterplots, with several outlying observations omitted, which gave the appearance of a more linear relationship than was present in the full dataset. Further complicating interpretation, the x-axis was labelled as log-transformed, although the plotted values corresponded to the untransformed data. This mislabelling made the reported model equations difficult to interpret, as the equations specified a log base 10 transformed predictor, whereas the transformation applied in the analyses was natural logarithms. | This paper was not inferentially reproducible, as all three models had linearity violations. In the Cycle Ergometer VO<sub>2</sub> model, these violations were largely driven by a single extremely influential observation. Linear (BIC = -61), quadratic (BIC = -87), and cubic (BIC = -215) specifications were examined; although the quadratic and cubic models showed substantial BIC reductions, none provided an adequate fit. The cubic model appeared to be dominated by extreme observations, while the linear and quadratic models were pulled downward at the upper end of the predictor range. After removal of the influential observation, the linear model remained a poor fit (BIC = -193), whereas the quadratic model fit best (BIC = -230). Log transformation improved the visual alignment between fitted values and the data; however, high-leverage observations continued to have a disproportionate influence on the OLS slope. Robust regression was therefore applied to down-weight these observations, producing a fitted relationship that more closely reflected the central trend. The remaining two models did not exhibit extreme outliers but showed violations of linearity, with linear terms failing to adequately characterise the associations. Both models were therefore refitted with log-transformed predictor and outcome variables. The log-log specification provided an adequate final model for the second analysis. In contrast, the third model continued to display residual curvature, and a natural cubic spline with three degrees of freedom was adopted as the final specification to allow modest curvature while limiting model complexity. Further modelling was constrained by the absence of participant-level identifiers. Without these identifiers, it is not possible to fit random-effects models or to formally assess whether the observed patterns reflect within-participant clustering rather than any underlying association. |

Gross violations of normality often suggested that an alternative outcome distribution would be more appropriate. The Gaussian (normal) distribution provided an adequate fit for most models (28) whereas a Tweedie distribution provided a better fit for three models and an Inverse Gaussian model for one. Linearity violations were identified in five papers, affecting 8 models. Further evaluation of model fit (AIC and BIC) and visual inspection of the data indicated that five of these models exhibited clear non-linearity.

Effect size comparisons between the reproduced and sensitivity results were restricted to directly comparable models, excluding those with specification differences (e.g., linear vs non-linear). Standardized regression coefficients were evaluated using percentage differences in estimates and changes in the confidence interval range, with interpretation primarily focused on the confidence interval range as an indicator of changes in precision rather than on changes in the mean effect alone. Across 97 coefficient ranges evaluated, 30 (31%) exceeded the 10% change threshold. When applying absolute thresholds based on standardized differences, 12 (12%) exceeded a change of 0.1 and 2 (2%) exceeded a change of 0.2. At the paper level, a paper was classified as exceeding a threshold if any coefficient range exceeded it. Nine of 11 (82%) papers exceeded the 10% threshold, while 5 (45%) exceeded the 0.1 threshold and 2 (18%) exceeded the 0.2 threshold, indicating a variation in classification depending on the threshold applied. Although coefficient thresholds were only one component of the reproducibility decision, a standardised difference of 0.1 was used as the threshold for determining reproducibility in this study.

In contrast to changes in confidence interval width, shifts in statistical significance were uncommon. Of the 97 p-values assessed, we observed only one (1%) shift in significance classification (from significant to non-significant or vice versa), with 40 (41%) remaining statistically significant and 56 (58%) remaining non-significant. Despite this relative stability in significance classification, a substantial proportion of models exceeded thresholds for changes in confidence interval width or demonstrated model assumption violations (e.g., linearity or independence) and were classified as not inferentially reproducible.

## Discussion

Despite the rapid growth in the analysis of health data, inferential reproducibility, the extent to which independent analysts draw similar conclusions from the same data, remains rarely assessed [3]. Our study highlights the practical challenges of evaluating this form of reproducibility, including incomplete reporting of statistical methods, vague model specifications, and limited information on key assumptions [20]. For example, some studies used repeated measures designs, yet the author-supplied datasets lacked participant identifiers, making it impossible to assess for the effect of clustering. This reflects a broader challenge of inferential reproducibility, which requires evaluating a series of complex modelling decisions that should be guided by the study design but are often poorly documented, making the process labour intensive.

Our results indicate that inferential reproducibility is often compromised when model assumptions are violated. In many cases, confidence intervals around estimates changed substantially, raising concerns about the reliability of the reported effects. Given that such findings may inform decisions about which health treatments are incorporated into standard practice, we recommend that studies making claims with potential clinical or policy implications be subject to inferential reproducibility assessments. Others have argued that reproducibility efforts should be strategically focused on high-impact studies, such as those with over 250 citations, to ensure that widely disseminated findings are trustworthy and to direct limited resources toward reproducibility [45]. However, this may be too late as by the time papers are cited this much, they are entrenched in the field; it may be better to focus on highly cited papers in the first year, e.g. 10 to 20. While awareness of reproducibility is growing, meaningful progress will require not only stronger reporting standards, targeted investment, and clearer incentives from journals and funders [46, 47], but also improved training in statistical reasoning and greater emphasis on evaluating model assumptions. These elements are essential to support researchers in making informed modelling decisions and producing reproducible, valid inferences.

Although automation is often proposed as a way to scale reproducibility checks, its feasibility depends on the aspect of reproducibility being assessed [48]. For inferential reproducibility, many modelling decisions require context-specific judgment that cannot be reliably captured through generic rules. While automation may help flag major assumption violations, such as extreme non-normality or heteroscedasticity, other decisions, such as how to address non-normality or whether to include non-linear terms, should not be based solely on the data, but guided by the study design and subject-matter considerations. These judgments require a clear understanding of both the model and the data. For this reason, any automated approach must be supplemented by expert human review, which can interpret assumptions in context and determine whether the chosen methods are appropriate.

### Comparison to Other Studies

A well-known case highlighting the consequences of inferential failure comes from economics. Reinhart and Rogoff [49] reported that countries with high debt-to-GDP ratios experienced lower economic growth, a finding that influenced global austerity policies. A spreadsheet error later uncovered by a graduate student revealed major flaws in the analysis [50]. These austerity measures led to reduced public spending in several countries, including cuts to health and social care budgets. In the UK, for example, such cuts were linked to excess deaths and widening health inequalities [51]. This case illustrates how analytical errors can shape policy with real-world health consequences, reinforcing the importance of reproducibility in evidence-based decision-making.

It is important to note that our study focused on statistical assumptions and did not consider variable selection, which can also affect reproducibility. This is clearly demonstrated by Silberzahn et al. [52], who asked 29 research teams to analyse the same dataset and research question, these types of studies are known as “*Many Analyst*” projects. Despite using the same data, teams applied 21 different covariate sets and a range of analytic methods, including linear regression, multilevel models, and Bayesian approaches, producing a large range in effect sizes. While 69% of teams reported statistically significant results, their conclusions varied, and choices regarding model type and handling of non-independence (e.g., variance components, clustered standard errors, or fixed effects) influenced the outcomes. The authors found that statistical expertise did not fully account for this variability, concluding that even well-intentioned analysts may reach different conclusions due to methodological flexibility in complex datasets.

Another *Many Analyst* project conducted by Gould et al. [53] showed how analytical decisions can substantially influence research findings, even when using the same data. In their study, 174 analyst teams analysed two ecological datasets on blue tit nestling growth and Eucalyptus seedling recruitment. The resulting effect sizes varied widely, with some teams finding strong effects, others near zero, and some even in the opposite direction. These findings further highlight how different analytical choices can lead to markedly different conclusions.

These many-analyst studies, like our findings, highlight that reproducibility is not just about data access but also how analytical decisions, such as model structure and handling of assumptions, can substantially influence conclusions [54]. While our study focused on inferential reproducibility, others have argued that reproducibility alone is insufficient. Munafo and Smith [55] advocate for triangulation using multiple, independent approaches to answer the same question, since replication can still reinforce flawed assumptions [56]. Although debate continues about the scale of the reproducibility “crisis,” there is broad agreement that improving methodological rigour and statistical reasoning is essential [57]. Fanelli [58] proposes reframing the issue as an opportunity for researchers to take greater responsibility for research quality.

### Interpreting Inferential Reproducibility: Beyond Statistical Significance

Our study contributes to the limited literature on inferential reproducibility by examining whether violations of linear regression assumptions influence study conclusions. We identified a number of papers in which linear regression was applied inappropriately, resulting in confidence intervals that were frequently too narrow. This underestimation of uncertainty suggests reduced precision and overstated confidence in the reported effects. In several cases, appropriately addressing these violations required fitting alternative models, such as replacing a linear model with a Tweedie or Inverse Gaussian model. A likely contributor to poor inferential reproducibility is limited understanding of model assumptions. Many researchers either do not formally assess assumptions or rely on statistical tests without recognising their limitations [9]. In our previous work, on the larger random sample of 95 papers, we found that the most common misconception was assessing normality of the outcome rather than the residuals [14]. Of the 28 author teams who reported checking normality, only five evaluated residuals, highlighting a substantial gap between theoretical knowledge and applied practice and reflecting persistent misunderstandings about the role and interpretation of regression assumptions.

One example from the current study where a violation of normality should have prompted reconsideration of the outcome distribution is found in paper 22. While two of the three models showed only mild deviations from normality of residuals, one showed a substantial deviation. Inspection of the outcome data revealed that nearly a quarter of the values were zero. Several modelling approaches are available for handling zero-inflated data. Two types of models address structural zeros, while a third treats zeros as part of a continuous mixture [23]. For instance, imagine a study measuring post-treatment quality-of-life scores on a 0–100 scale, where zero represents no impairment. Thirty percent of patients report a score of zero, while the remaining 70% have scores between 1 and 100. This pattern fits a Tweedie distribution, a flexible family that includes the Poisson and Gamma distributions as special cases and accommodates a mean–variance relationship in which the variance increases with the mean. Alternatively, a hurdle model is appropriate when zeros arise solely from a structural mechanism, for example, when a patient did not experience the condition being assessed and therefore could not have an impairment. A zero-inflated scenario occurs when both structural zeros (patients who could not have impairment) and sampling zeros (patients who could have impairment but reported none) are present in the same dataset. The complexity of model choice and interpretation highlights the importance of consulting an experienced statistician, as papers involving statisticians tend to have fewer errors, including inappropriate test selection [59].

Assessing whether model assumptions are valid is often more similar to a mixed-methods approach than a purely quantitative process. As in qualitative research, where triangulation of multiple data sources can assess whether they tell a consistent story, assumption checking should integrate both quantitative indicators and contextual understanding. For example, knowledge of the study design is essential when assessing independence. Quantitatively, the model can be examined for the level of correlation between measurements using the intra-class correlation coefficient (ICC); the AIC can inform model complexity; and effect sizes and confidence intervals can be compared across models to assess changes. While one of these may be the dominant consideration in some cases, a full evaluation typically requires combining all sources of information to make an informed decision. For instance, when the ICC is relatively low, researchers might still choose to use a linear mixed model rather than a simple linear regression to remain consistent with the study design. However, a high ICC indicates that simple linear regression is inappropriate. We observed this in paper 16, where both models assessed had high ICCs (0.26 and 0.35), indicating substantial within-cluster correlation.

In our study, p-value classifications were relatively stable, with few changes from statistically significant to non-significant and vice versa. However, this finding should be interpreted in context and cannot be directly compared to studies reporting inconsistencies in p-values [60], as our analyses were restricted to models that had already been found to be computationally reproducible. Some may argue that if statistical significance remains unchanged, there is little cause for concern. Yet statistical significance is a poor arbiter of importance. Small studies often remain non-significant despite meaningful shifts in effect magnitude, whereas large studies can demonstrate statistical significance even when changes are trivial or clinically irrelevant. Our assessment showed that many authors framed their conclusions primarily in terms of statistical significance rather than effect magnitude or precision [15]. Although significance categories were relatively stable, confidence interval widths were frequently not reproducible. This indicates that unchanged statistical significance does not imply inferential stability. Precision is particularly important when estimating clinically relevant change, as the width of the confidence interval helps define the range of plausible effects and therefore the degree of certainty when weighing potential benefits against risks for new treatments.

To evaluate changes in effect size, we examined three thresholds. A threshold of 10% change in estimates flagged many inconsequential differences, and we do not recommend this approach for assessing inferential reproducibility. In contrast, absolute thresholds of 0.1 and 0.2 for changes in standardized regression coefficients performed more appropriately. The lower threshold (0.1) was more consistent with cases in which model violations were present. For example, in one instance with an ICC of 0.26, the change in the range of the standardized confidence interval was approximately 0.15. A simulation study to evaluate the utility of these thresholds under controlled conditions would be a valuable next step.

### Challenges in Assumption Checking

Our framework does not require nearly identical estimates or consistency across every reasonable statistical approach. Rather, it assesses whether the original inference remains credible when the data are analysed using an alternative approach that addresses an identified problem such as an assumption violation. Although the direction of association and dichotomised statistical significance were often unchanged, these features alone do not establish inferential reproducibility. Ordinary least squares (OLS) is the standard method used to estimate coefficients in linear regression. The coefficients can still be calculated when model assumptions are not satisfied, but the validity of the estimates and conventional standard errors, confidence intervals, and p-values depends on the relevant assumptions. An unchanged coefficient direction or significance category therefore does not demonstrate that the estimated effect, its uncertainty, or any resulting interpretation is reliable.

Regression assumptions are not interchangeable or additive. Several minor departures may have little practical effect, whereas a single substantial violation, such as failing to account for repeated measurements or fitting a linear model to an outcome with an incompatible mean–variance relationship, may be sufficient to undermine the estimated uncertainty and resulting inference. The final assessment was therefore expressed as a binary (Yes/No) classification of whether the original inference remained credible, rather than as a graded or additive score that could imply that a serious violation can be offset by satisfactory performance in unrelated areas.

An important consideration in evaluating model assumptions is that the process requires judgment and visualisation, rather than relying solely on strict, hypothesis-testing-based rules, which can sometimes be misleading [61]. Even statisticians may reasonably disagree on the best approach. Multi-analyst studies have demonstrated how different analytic decisions can yield varying results [53, 52]. It is also important to note that residuals are estimates of the true errors [62]. Overly flexible approaches, such as fitting Generalized Additive Models (GAMs) to residuals can overfit noise, mistaking random variation for structure [63, 64]. While minor violations of assumptions may not always justify alternative models, we observed substantial violations in several papers in which linear regression was inappropriate yet used without question.

These challenges are compounded by the way statistics is commonly taught. Introductory courses often present methods such as t-tests, ANOVA, and linear regression in a simplified, rule-based manner, typically applied to clean, “well-behaved” data [65]. These models are rarely introduced as part of a unified framework, namely the General Linear Model (GLM), leading students to view them as unrelated procedures rather than variations of the same underlying structure [14]. Measures such as R^2^ are frequently taught as key indicators of model quality, despite offering limited insight into model appropriateness or reliability. Broader tools for model assessment, such as the Akaike Information Criterion (AIC), may be less often introduced, leaving researchers with a limited understanding of how to evaluate fit or compare models. Moreover, violations of statistical assumptions are often treated as technical checklist items rather than potential indicators of model misspecification. For example, apparent non-normality of residuals may reflect omitted variables or an incorrectly specified relationship between variables rather than a problem with the outcome distribution.

Similarly, outlier handling in some textbooks and basic statistics courses is often taught using simple cut-offs (e.g., three standard deviations from the mean), rather than through influence diagnostics such as Cook’s distance, DFFITS, or DFBETAS. While these measures focus on changes in regression coefficients, diagnostics that assess the precision of estimates, such as COVRATIO, which evaluates the inflation of standard errors and the potential widening of confidence intervals, are often neglected, leaving applied researchers without a complete picture of how outliers affect inference [26]. Many textbooks also provide conflicting advice on whether to exclude outliers, often without evaluating the appropriateness of removal based on influence or its impact on model validity [66].

Although excluding outliers without strong justification is generally inappropriate, influential observations should be explicitly examined in sensitivity analyses to assess their impact on estimates and inference. Approaches such as robust regression and bootstrapping are commonly used in this context. Robust regression downweights influential observations during model fitting, thereby reducing their impact on coefficient estimates and associated inference. Bootstrapping can reduce reliance on strict distributional assumptions by resampling the observed data; however, it assumes that the fitted model is correctly specified and that influential observations are representative of the underlying data-generating process. Neither approach addresses fundamental model misspecification, such as incorrectly modelling the relationship between variables.

When influential observations were identified, their impact was evaluated to determine whether they distorted parameter estimates or reflected broader model-assumption violations. In paper 46, linearity and outlier concerns were addressed by log-transforming both the predictor and outcome. However, high-leverage observations continued to exert disproportionate influence on the fitted slope. Robust regression was therefore applied to the log–log model to down-weight these observations during estimation, producing coefficients that more closely reflected the central trend in the data. A similar issue arose in paper 32, where log-transformation addressed departures from linearity and substantially reduced the influence of extreme observations. However, residual heteroscedasticity persisted, indicating a violation of the constant-variance assumption. Robust standard errors were therefore applied to the log–log model to obtain valid inference without altering the point estimates. These examples highlight the distinct roles of robust methods: robust regression is appropriate when influential observations distort parameter estimates, whereas robust standard errors are appropriate when variance assumptions are violated but the model is otherwise adequate.

Papers 32 and 46 also illustrate why models should not be selected solely on the basis of AIC or BIC Table 2. Although these criteria compare the relative fit of candidate models while penalising model complexity, the model with the lowest value is not necessarily the most scientifically plausible. In these examples, polynomial terms initially appeared to improve model fit, but graphical assessment showed that the apparent relationships were strongly influenced by extreme observations. A higher-order polynomial may similarly improve AIC or BIC by closely following features specific to the observed dataset while representing a relationship that is difficult to justify substantively or unlikely to generalise. Information criteria should therefore be interpreted alongside graphical assessment, residual diagnostics, theoretical plausibility, and the influence of individual observations.

Some argue that many authors do check model assumptions but, due to space constraints in journals, do not report the details or even mention that checks were conducted [67]. To our knowledge, no empirical research has formally tested this claim. Our findings suggest that the proportion of papers clearly addressing assumptions is low. While some evidence suggests that a few authors assessed assumption violations, these violations were rarely reported in detail. For example, one paper stated that assumptions were checked and found to have only mild violations that did not substantially affect results. However, many of the papers we assessed for inferential reproducibility reported only limited, and often incorrect, assumption checks, despite major violations that substantially altered the precision of the reported effects.

Issues of assumption checking also intersect with broader concerns about modelling choices and reproducibility, where software defaults may shape analytic decisions. One example we observed is from paper 22, in which ethnicity was incorrectly modelled as a continuous variable. In SPSS, unless categorical variables are explicitly dummy coded, they are treated as continuous in linear regression models. Similar risks arise when stepwise regression is applied to dummy-coded categorical variables and interaction terms, where software may treat individual terms as independent. A possible example is from paper 38, where substantial effort was devoted to testing assumptions of their interaction model; yet backward stepwise regression was subsequently used, resulting in the elimination of main effects involved in the interaction and components of categorical variables. SPSS also includes a function called Univariate General Linear Model, which performs linear regression within the broader General Linear Model framework. This function automatically dummy-codes categorical variables, which researchers may not realise given its name and setup, although it does not perform variable selection. These examples highlight a key point: understanding your software’s default settings is essential. Seemingly minor choices, such as drop-down selections or default parameterisations, can substantially change the interpretation of results. Without adequate statistical training, researchers risk producing results that are computationally reproducible but inferentially misleading.

Our findings highlight the need for better statistical education that reflects the complexity and judgment involved in modelling. Qualitative evidence suggests that researchers often interpret regression coefficients through context dependent reasoning rather than as purely mechanical statistical outputs [68], reinforcing the importance of conceptual understanding over rule based application. Some authors in our sample made genuine efforts to satisfy regression assumptions, but this sometimes led to models that were difficult to interpret and overemphasised statistical significance. For example, transformations were sometimes applied and not back-transformed, limiting interpretability on the original scale. In other cases, transformations appeared to be motivated primarily by a desire to “pass” normality tests, even when deviations were unlikely to be practically meaningful. Teaching statistics within a broader modelling framework, one that emphasises critical thinking over rigid rules, would better prepare researchers to apply methods appropriately, consider alternatives such as bootstrapping or other distributions, and prioritise interpretability and practical relevance over arbitrary thresholds.

### General Recommendations for Improving Inferential Reproducibility

- Consider the underlying data-generating mechanism (e.g. continuous, count, or proportions) when selecting analytical approaches. Provide exploratory visualisations in the Supplementary Information, such as scatterplots, boxplots, or other appropriate plots, to illustrate key relationships between variables included in the analyses.
- Clearly describe the study design, explicitly stating whether the study is observational or experimental, whether randomisation was used, and whether there is temporal ordering, such as in a baseline and follow-up design.
- Provide a unique observation identifier (ID) in the dataset to ensure each individual is uniquely and consistently identified.
- Consider whether the study design includes clustering (e.g. by site, school, hospital, or family). If clustering is present, include the relevant clustering variables in the dataset.
- Clearly describe and justify the modelling strategy, including the rationale for inclusion of confounders, mediators, and interaction terms, and whether a formal model-building framework was used.
- If a formal modelling process was used, the variable selection and/or removal procedures should be described in detail, including all decision thresholds and criteria. Authors should also report the number and type of candidate variables initially considered for inclusion, rather than only listing those retained in the final model.
- Evaluate the assumptions of linear regression models by examining residual diagnostics, identifying influential observations, and assessing multicollinearity among predictors. Where assumptions are violated, apply and report appropriate remedial methods.
- If model assumptions are violated, consider fitting alternative models that better reflect the underlying data structure (e.g. non-linear specifications or models with different distributional assumptions). Use appropriate model-fit statistics, such as R2, Akaike Information Criterion (AIC), or Bayesian Information Criterion (BIC), to compare competing models and justify the final model choice.
- Assess heteroscedasticity explicitly and, where present, consider using robust standard errors or alternative modelling strategies to ensure valid statistical inference. When systematic variance differences are evident (e.g., differing variances across groups or increasing variability with a predictor), weighted least squares may be considered with appropriate justification.
- Identify influential observations using appropriate influence diagnostics (e.g. Cook’s distance, DFBETAs, DFFITS, and covariance ratios), and evaluate their impact on coefficient estimates and statistical inference. Where influential observations materially affect results, conduct sensitivity analyses, such as applying robust regression methods to reduce their influence.
- Transformations may be used to address violations of linearity or homoscedasticity, but should be applied only where they meaningfully improve model adequacy while retaining interpretability. For log-transformed models, back-transformation is often required to present results on the original scale, such as geometric means or ratios (interpreted as percentage or fold changes). Researchers should consider whether a transformation is necessary and whether the resulting parameter estimates correspond to the effect of interest, recognising that transformations (e.g. log scale) change interpretation from additive differences to multiplicative or proportional effects.
- Bootstrapping can provide standard errors and confidence intervals without relying on the assumption that residuals are normally distributed. However, it does not address model misspecification, such as non-linearity. When bootstrapping is used, the resampling scheme (e.g., residual, case, wild, or cluster bootstrap), the number of resamples, and the model coefficients and confidence intervals should be clearly reported.
- Report the details of assumption checking in the Supplementary Information, including diagnostic plots and any sensitivity analyses undertaken.
- Provide a clear description of the extent and patterns of missing data, including the assumed missing data mechanism (e.g., missing completely at random, missing at random, or missing not at random). Authors should detail the analytical approach used to address missingness (e.g., complete case analysis, single or multiple imputation, inverse probability weighting), along with any sensitivity analyses conducted to assess the stability of results.
- Share code, underlying data, and a comprehensive data dictionary with a well-documented workflow to clarify modelling decisions and support inferential reproducibility.

### Limitations

While every effort was made to faithfully reproduce published analyses using the methods described in the original articles, evaluating statistical assumptions is inherently subjective. Contextual knowledge, often unavailable to external analysts, can influence judgments about the severity and relevance of assumption violations. Consequently, reasonable differences in interpretation and analytical decisions may arise among statisticians and researchers. In addition, implicit methodological details, such as data-handling procedures or variable coding, were often not fully reported, which may affect reproducibility. Accordingly, the individual classifications and overall inferential reproducibility estimate reflect what could be independently assessed from the published articles and shared materials; additional undocumented contextual information may have altered some assessments. Nonetheless, we maintain that research articles should provide sufficient detail for any qualified statistician to independently reproduce the analyses.

As this was an exploratory study, the methodology was refined as challenges were identified; the study protocol is available on GitHub [30]. Our results should be interpreted with caution, as we only assessed inferential reproducibility in papers that were computationally reproducible, a subset that may disproportionately represent methodologically skilled authors and was conducted using a relatively small number of papers. The primary author evaluated inferential reproducibility, with input from co-authors as required. Although our protocol initially specified a 10% change in coefficients as a threshold, this was often not meaningful when coefficients were close to zero. To address this, additional comparisons were made using cut-offs of 0.1 and 0.2 for changes in standardized regression coefficients. These pragmatic cut-offs were informed by the research team’s experience in judging whether small differences were substantively meaningful and by conventional benchmarks for small effect sizes. However, further research is needed to validate these thresholds.

Assumption violations were not considered consequential solely because they were present, as their effects depend on their nature, severity, and the context of the analysis. When a violation was identified, expert judgement was used to assess its potential importance, and sensitivity analyses were conducted to determine whether addressing it materially affected the estimates or resulting inferences. This process required judgement in selecting appropriate alternative models and determining whether observed differences were substantively meaningful. Despite these limitations, our findings are consistent with previous research documenting concerns about inferential reproducibility in scientific research [3] and limited or inconsistent reporting of model assumptions [69].

## Conclusions

Assessing inferential reproducibility is inherently complex, as it requires judgment about whether modelling choices and assumptions are appropriate for the study design and research context. While major issues, such as extreme non-normality or severe heteroscedasticity, are relatively straightforward to identify, more nuanced decisions, such as whether to include non-linear terms or account for zero inflation, require contextual understanding and cannot be reduced to simple rules without risking misinterpretation.

The limited reporting of assumption checks observed in the sample of papers we examined suggests that modelling decisions may not always be explicitly justified. In some instances, model specifications appeared consistent with default software settings rather than decisions clearly aligned with the research question. Linear regression models were frequently used without explicit assessment of key assumptions or consideration of whether it was the most appropriate modelling approach.

To help guide researchers, we have developed general recommendations for inferential reproducibility informed by issues identified across the studies examined. The recommendations draw directly on recurring problems observed in the reporting and interpretation of statistical analyses and are intended to provide practical guidance to improve the transparency and consistency of inference. We recommend using these recommendations when developing statistical analysis protocols, as they promote explicit, up-front consideration of modelling assumptions, analytical decisions, and planned sensitivity analyses, and revisiting them during the analysis stage to ensure these considerations are implemented and transparently reported. In our experience, introductory statistics courses are often taught in ways that encourage simplified responses to violations of assumptions, such as defaulting to nonparametric tests. This can leave researchers insufficiently prepared to manage the complexities of statistical modelling. We recommend introducing the foundational principle that “everything is a regression” [70], whereby t-tests, ANOVA, and linear regression are understood as special cases of the general linear model. Extending this framework to include link functions and Generalised Linear Models would help students develop a deeper understanding of modelling from the outset. Although it is not feasible to teach all statistical techniques in a single introductory course, conveying this broader conceptual framework is particularly important given that many health professionals complete only one formal course in statistics during their university degrees.

As analyses become more complex, researchers should be encouraged and supported to consult with qualified statisticians, ensuring that modelling choices are appropriate, assumptions are adequately evaluated, and results are valid and interpretable. However, in many hospital and health service contexts, access to this expertise is limited. This is particularly concerning given the potential consequences of low-quality studies in health settings, where inappropriate modelling or unchecked assumptions can lead to misleading conclusions. A stronger understanding of how decisions such as model parameterisation and distributional assumptions influence conclusions is essential for informed decision-making. Reproducibility is more than a technical exercise; analyses may computationally reproduce yet still produce misleading conclusions if they rely on flawed modelling choices.

While minor violations of statistical assumptions should be discussed in manuscripts and comment sections, situations involving gross violations, such as changes to substantive conclusions or the use of inappropriate models, warrant a formal correction. At present, however, practices such as sharing data or documenting model assumptions are often treated as optional or even risky, with the potential to result in negative professional consequences if errors are discovered [71]. A further barrier is the perception that acknowledging mistakes or issuing corrections damages a researcher’s and journals’s reputation. Journals and publishers could help reduce these barriers by embedding reproducibility into standard expectations and by making the correction process straightforward and routine. Institutions also have a responsibility to foster a culture in which correcting errors is expected and valued, positioning transparency and correction as markers of professional integrity rather than sources of stigma.

Journals also have a crucial role in strengthening peer review by engaging qualified statistical reviewers and enforcing compliance with data availability requirements. Some disciplines have adopted more structured editorial models to support reproducibility; for example, the American Economic Association employs a dedicated data editor to review replication materials prior to publication [72]. Similar mechanisms in health research could improve scrutiny of modelling decisions and enhance inferential reproducibility.

However, inferential reproducibility problems should not be attributed solely to gaps in individual researchers’ statistical knowledge. Publication incentives, including pressure to publish, and broader systemic pressures may limit the time, resources, and support available for careful model development, diagnostic assessment, and transparent reporting [73]. Limited peer-review capacity and inconsistent access to methodological expertise may further reduce opportunities for statistical problems to be identified before publication [74, 75].

Our findings reinforce ongoing calls for stronger methodological and statistical rigour in health research. While computational reproducibility is valuable, it is insufficient to ensure scientific reliability. Inferential reproducibility is essential for understanding the magnitude and precision of results and depends on appropriate modelling decisions, interpretation, and assessment of assumptions. Meaningful improvements will require improved statistical literacy, practical training in model diagnostics, and systemic changes that support high-quality research practices. Reproducibility must be understood not merely as the ability to rerun code, but as the capacity to justify, evaluate, and defend statistical decisions in a transparent and principled manner. Strengthening inferential reproducibility is therefore central to improving the credibility, interpretability, and practical value of health research.

## Data Availability

The data and a reproducible R Quarto file used to produce this paper, including tables, figures, and code, have been stored in a GitHub repository and can be cited using the Zenodo doi.

https://github.com/Lee-V-Jones/Reproducibility

https://doi.org/10.5281/zenodo.19448969

## Acknowledgements

We acknowledge Adrian llich for providing bioinformatics support and all the statisticians, including those acknowledged by name and others who chose to remain anonymous, who generously contributed their time to reviewing papers for this study, including: Ingrid Aulike, Peter Baker, Brigid Betz-Stablein, Enrique Bustamante, Taya Collyer, Susanna Cramb, Alanah Cronin, Laura Delaney, Zoe Dettrick, Eralda Gjika Dhamo, Des FitzGerald, Peter Geelan-Small, Edward Gosden, Alison Griffin, Jenine Harris, Cameron Hurst, Kyle James, Helen Johnson, Jessica Kasza, Karen Lamb, Stacey Llewellyn, James Martin, Miranda Mortlock, Satomi Okano, Alan Rigby, Michael Steele, Megan Steele, Jacqueline Thompson, Simon Turner, Michael Waller, Kevin Wang, Jace Warren, Natasha Weaver, Lachlan Webb, and Janet Williams.

